# Cohort profile: The WHO Child Mortality Risk Stratification Multi-Country Pooled Cohort (WHO-CMRS) to identify predictors of mortality through early childhood

**DOI:** 10.1101/2024.03.06.24303859

**Authors:** Catherine Schwinger, Siri Kaldenbach, James A. Berkley, Judd L. Walson, Alemayehu Argaw, Ranadip Chowdhury, Tor A. Strand, Nigel Rollins, the WHO Risk Stratification Working Group (WHO-RSWG)

**Affiliations:** Centre for International Health, University of Bergen, Bergen, Norway; Department of Research, Innlandet Hospital Trust, Lillehammer, Norway; Kenya Medical Research Institute (KEMRI)/ Wellcome Trust Research Programme, Kilifi, Kenya; Centre for Tropical Medicine and Global Health, Nuffield Department of Clinical Medicine, University of Oxford, Oxford, UK; Department of International Health, Bloomberg School of Public Health, Johns Hopkins University, Baltimore, USA; Department of Food Technology, Safety and Health, Faculty of Bioscience Engineering, Ghent University, Ghent, Belgium; Centre for Health Research and Development, Society for Applied Studies, New Delhi, India; Department of Maternal, Newborn, Child and Adolescent Health and Ageing, WHO, Geneva, Switzerland

**Keywords:** Age-specific death rate, anthropometry, morbidity, low birthweight, preterm birth, breastfeeding, LMIC

## Abstract

**Purpose:** To provide details of a pooled dataset that will be used to estimate absolute and relative mortality risks and other outcomes among children less than 59 months of age and the predictive performance of common risk exposures, both individually and in combination.

**Participants:** Children from birth to five years of age recruited at health facilities or community settings into 33 longitudinal observational or intervention studies in 18 low-and middle-income countries.

**Findings to date:** The dataset includes 75,287 children with a median age of 3 months (IQR 1, 12) at first measurement. In the pooled sample, 2,805 (3.7%) of the study children died. Data on birthweight was recorded in 18 studies, and gestational age in 13 studies. Among these, 14% of the included children were reported with low birthweight and 14% preterm birth. At first measurement, 33% of the children were stunted, 24% were wasted, and 35% underweight. 13% and 7% of caregivers reported that their child had acute diarrhoea or acute lower respiratory tract infection before the study visit, respectively. The proportion of children being breastfed at any study visit decreased from 99% at age <6 months to 77% in the age group 12-23 months. Child characteristics differed considerably between studies in the community and health care settings. The median study period was 15 months (IQR 7.6 to 18.4 months).

**Future plans:** The WHO Child Mortality Risk Stratification Multi-Country Pooled Cohort (WHO-CMRS) provides a large dataset including child, parental, and household characteristics from a diverse range of geographical, community and health system settings; planned analyses will examine knowledge gaps with the aim of informing global guidelines and their derivatives such as clinical management tools and implementation guidance, and to inform future research agendas.

**STRENGTHS AND LIMITATIONS:** *Strengths:*

– A large dataset that integrates diverse geographical areas in LMICs, with children recruited from community and health system settings.
– Prospective data collection and availability of key time-varying exposures, such as nutritional status, common childhood morbidities, and feeding practices and non-time varying exposures, such as birth characteristics and socio-demographics.
– Ability to estimate age-specific absolute mortality risks from different exposure domains and combinations thereof.

*Limitations:*

– Non-systematic selection of included datasets.
– Heterogeneity of inclusion criteria, exposures collected and follow-up schedules across studies.

## INTRODUCTION

Sustainable Development Goal (SDG) 3 aims to end preventable deaths of newborns and children under five years of age by 2030. Although the world has seen remarkable progress in reducing deaths in children under 5 years, with a 59% reduction since 1990, preventable child mortality is still high. The rate of decline has also decreased since 2015. In 2021, an estimated 5 million children under the age of five died of largely preventable or treatable causes (1, 2). Furthermore, 53 countries must double or triple their current pace in reducing the under-five mortality rates to meet this SDG. There is an urgent need to identify and support improvements in strategies to protect child survival.

Communicable diseases, together with preterm birth, remain the leading (direct) causes of death in children (1, 2), with pneumonia being the single most significant infectious cause of death in children worldwide (3–5). Undernutrition is estimated to be associated with up to 50% of all deaths in under-five children (6), and the risk of death increases with greater degrees of anthropometric deficit (7). Nevertheless, there is longstanding debate within the nutrition research and practice communities surrounding the most appropriate anthropometric measures to identify the risk of death (8, 9), especially for infants under six months of age (10). Low birthweight (11, 12), preterm birth (13–15), and sub-optimal breastfeeding practices (16) as well as maternal nutrition and vital status (17) are also well-established risk factors. Focusing on single risk domains such as anthropometry or disease severity may overly simplify the risks for an individual child who may present with more than one disease and/or underlying conditions (18), and who may have additional environmental, social and economic risk factors not captured by clinical risk assessments (19).

The concept of risk stratification is integral to WHO care pathways, such as the Integrated Management of Childhood Illness (IMCI) or Integrated Community Case Management. However, these clinical tools largely focus on the severity of the presenting signs, such as dehydration or fast breathing or chest indrawing. Although some evidence is emerging that more systematically addresses the interaction of undernutrition and morbidity on mortality (20–23), there is limited literature exploring the general concept of risk stratification at the individual level for vulnerable children (24, 25). Identifying children at the highest risk of death, especially in resource-constrained settings, may enable formal health systems to target these groups more effectively and improve resource allocation and programmatic focus. In addition, identifying children at much lower risk of death may provide the opportunity to redirect limited resources to achieve maximal impact. We pooled multiple datasets to establish a large database that integrates individual-level data from a range of geographical, community and health system settings in LMICs. Analyses are in progress to estimate absolute and relative mortality risks in early childhood and predictive performance of common risk exposures, both individually and in combination. This initiative may provide evidence to inform global guidelines, clinical management tools and future research agendas benefitting infants and children at the highest risk of adverse outcomes. Here we describe the process of bringing the pooled dataset together as well as the main characteristics of the studies and children whose data are included. These will inform future analysis approaches.

### COHORT DESCRIPTION

#### Selection of studies

Between August and November 2021, WHO convened the Risk Stratification Working Group (RSWG) and invited research investigators to consider specific research questions and analytical approaches for a set of risk stratification analyses related to child mortality. Researchers were invited based on known interest and having published relevant data. Other researchers requested to participate and actively contributed to the technical discussions. A list of RSWG members can be found in Table S1. Collectively, it was agreed that an individual-level pooled analysis of prospective cohort data would provide the highest quality evidence, enabling harmonization of an analytical approach, inclusion of smaller datasets, as well as flexibility for interaction and sensitivity analyses.

Datasets that could contribute to the analyses were identified following active outreach by all RSWG members within their various networks, emphasizing an inclusive approach.

In February 2022, all identified eligible study teams interested in sharing their data with the RSWG were asked to complete a Data Pooling Agreement. The agreement outlined conditions by which the Principal Investigators or representatives of host institutions of specific studies would share anonymized data with WHO for inclusion in the joint analyses, with the WHO being responsible for the overall coordination and additional technical support being provided by the University of Bergen. In total, 27 agreements were signed granting permission to pool 35 separate datasets. The WHO Ethics Review Committee granted an exemption in March 2022 with protocol ID: ERC.0003745.

The following inclusion criteria were set for datasets to be included in the pooled database:

- The dataset included a minimum set of variables defined as “required”, i.e., mortality, age, sex, and weight. Data on other anthropometric measures, and exposures within other risk domains including morbidity, maternal health and nutrition status, pregnancy outcomes, and clinical signs and symptoms were requested, but not required for inclusion in the dataset.
- The study included data on children <60 months of age.
- The study was conducted in a low-or low-middle income country (World Bank definition) (26).
- The study followed individual children longitudinally.
- An adequate description of the study population including the sampling and recruitment strategy (e.g., random vs. convenience sampling, self-presentation vs. active community screening), inclusion and exclusion criteria, and assessment procedures were available.
- Date of enrolment of participants from January 2000 onward.

Datasets were excluded if they were cross-sectional or did not include all essential data i.e. mortality, age, sex, and weight.

#### Data sources in the pooled dataset

Up until August 2023 and following the exclusion of two datasets, 33 datasets had been included in the pooled database (Figure 1). Individual studies contributing to the pooled database were conducted in 18 different countries in five of the six WHO Regions: six in South-East Asia, one in the Eastern Mediterranean region, 24 in sub-Saharan Africa, one in the Western Pacific region, and one in Latin America. One study was conducted in more than one world region (Study 30, Table 1). Of the studies included, 21 were randomized controlled trials (RCTs), and 12 were observational cohort studies. Most RCTs (16 studies) assessed nutritional interventions such as multivitamins or specially formulated and nutrient dense foods with or without additional educational or biomedical elements. Included studies are described in Table S2.

**Figure 1:**
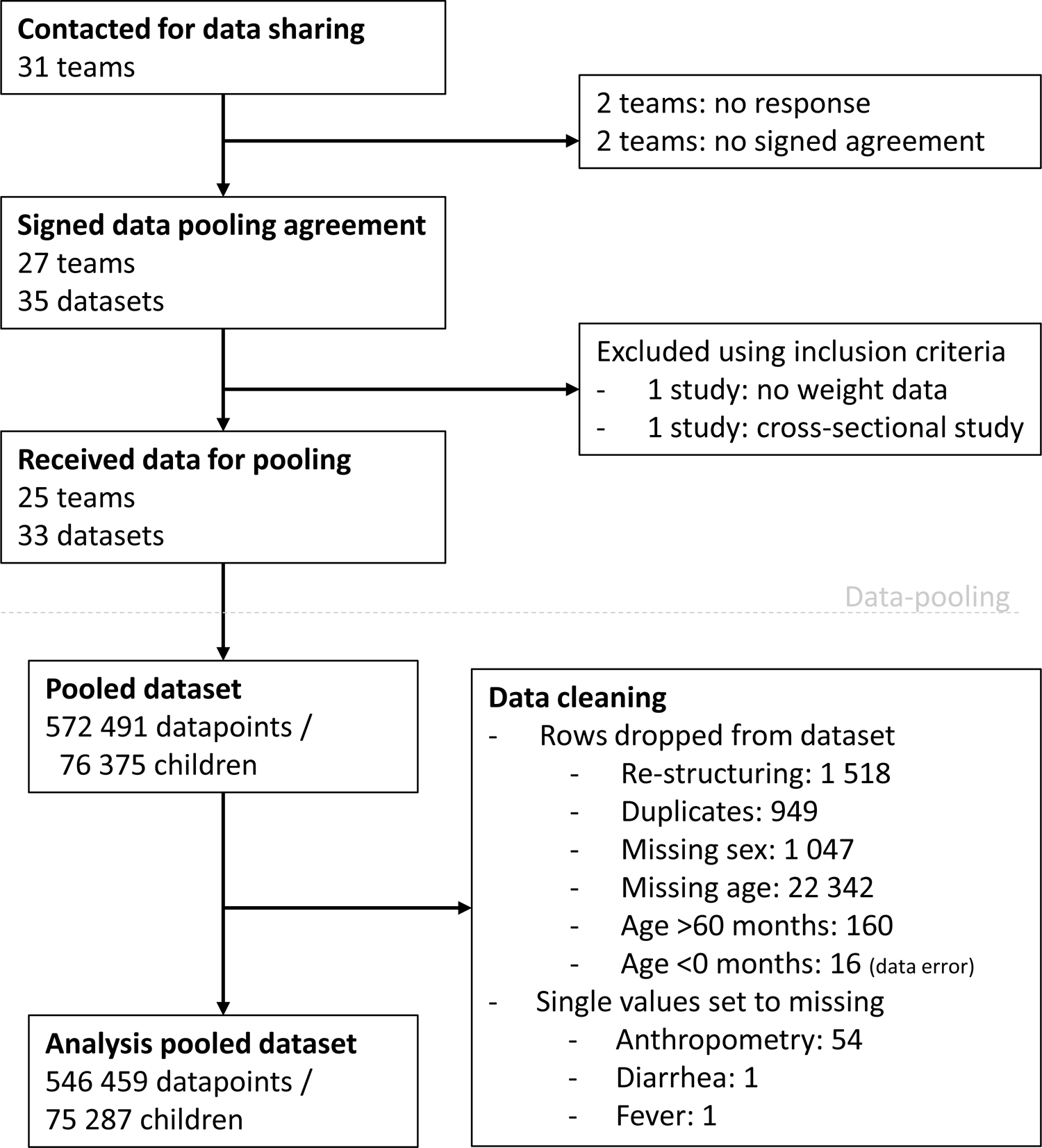
Overview of the data included in the WHO Child Mortality Risk Stratification Multi-Country Pooled Cohort (WHO-CMRS)

**Table 1:**
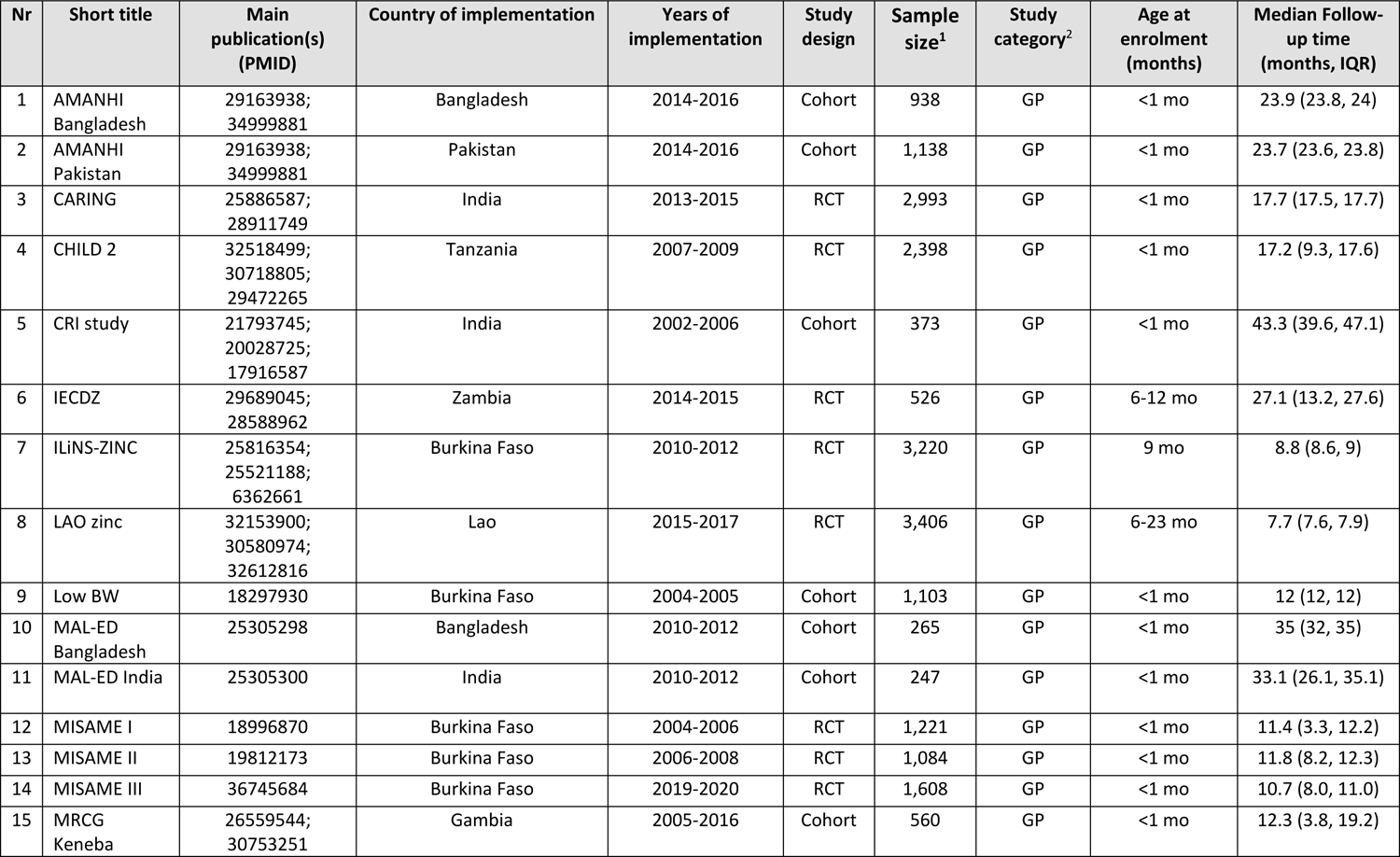
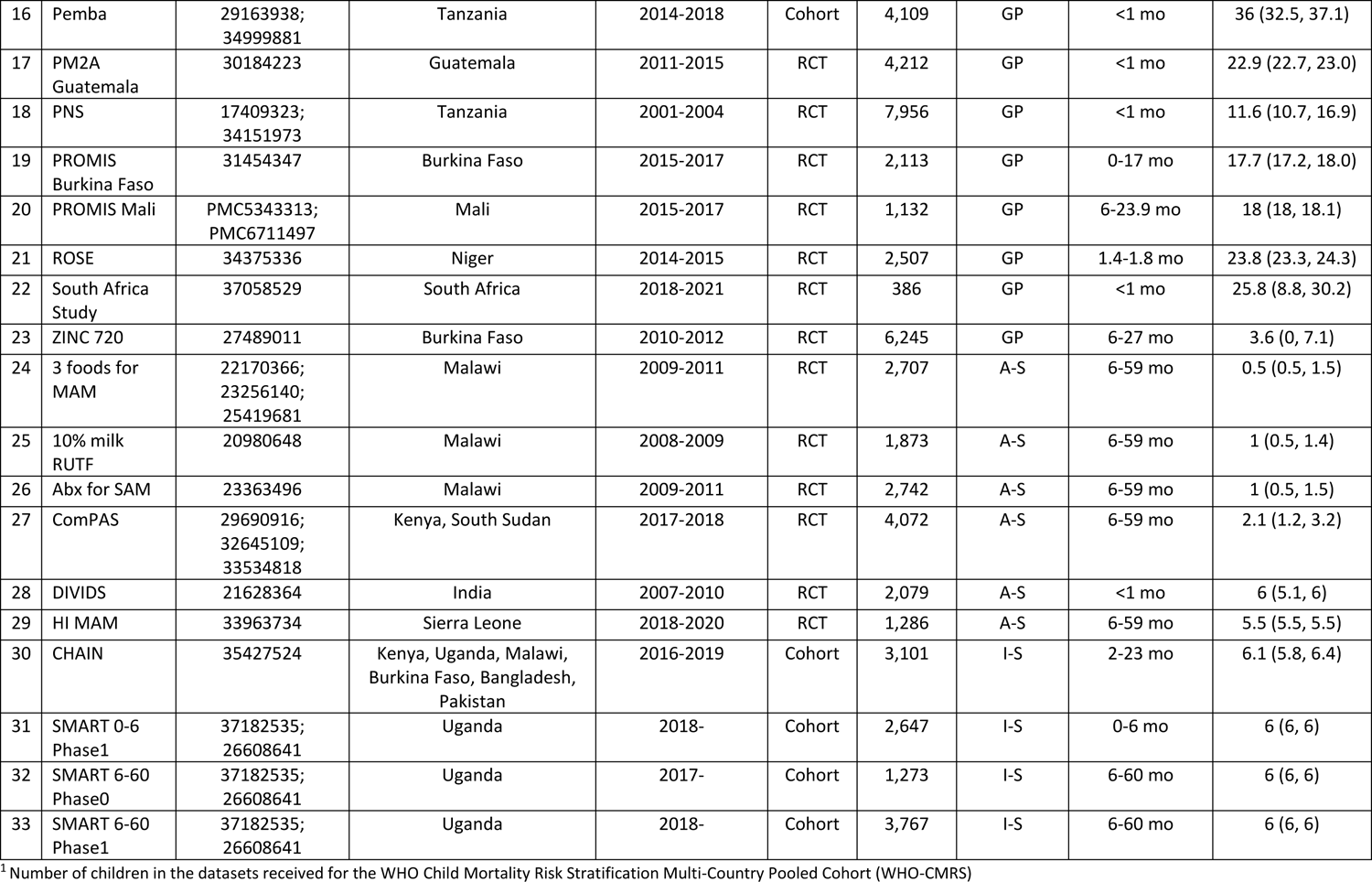
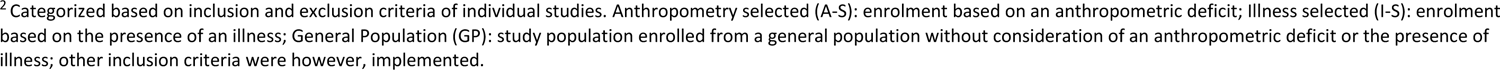
Brief description of studies included in the WHO Child Mortality Risk Stratification Multi-Country Pooled Cohort (WHO-CMRS)

| Nr | Short title | Main publication(s) (PMID) | Country of implementation | Years of implementation | Study design | Sample size<sup>1</sup> | Study category<sup>2</sup> | Age at enrolment (months) | Median Follow-up time (months, IQR) |
| --- | --- | --- | --- | --- | --- | --- | --- | --- | --- |
| 1 | AMANHI Bangladesh | 29163938;<br>34999881 | Bangladesh | 2014-2016 | Cohort | 938 | GP | <1 mo | 23.9 (23.8, 24) |
| 2 | AMANHI Pakistan | 29163938;<br>34999881 | Pakistan | 2014-2016 | Cohort | 1,138 | GP | <1 mo | 23.7 (23.6, 23.8) |
| 3 | CARING | 25886587;<br>28911749 | India | 2013-2015 | RCT | 2,993 | GP | <1 mo | 17.7 (17.5, 17.7) |
| 4 | CHILD 2 | 32518499;<br>30718805;<br>29472265 | Tanzania | 2007-2009 | RCT | 2,398 | GP | <1 mo | 17.2 (9.3, 17.6) |
| 5 | CRI study | 21793745;<br>20028725;<br>17916587 | India | 2002-2006 | Cohort | 373 | GP | <1 mo | 43.3 (39.6, 47.1) |
| 6 | IECDZ | 29689045;<br>28588962 | Zambia | 2014-2015 | RCT | 526 | GP | 6-12 mo | 27.1 (13.2, 27.6) |
| 7 | ILiNS-ZINC | 25816354;<br>25521188;<br>6362661 | Burkina Faso | 2010-2012 | RCT | 3,220 | GP | 9 mo | 8.8 (8.6, 9) |
| 8 | LAO zinc | 32153900;<br>30580974;<br>32612816 | Lao | 2015-2017 | RCT | 3,406 | GP | 6-23 mo | 7.7 (7.6, 7.9) |
| 9 | Low BW | 18297930 | Burkina Faso | 2004-2005 | Cohort | 1,103 | GP | <1 mo | 12 (12, 12) |
| 10 | MAL-ED Bangladesh | 25305298 | Bangladesh | 2010-2012 | Cohort | 265 | GP | <1 mo | 35 (32, 35) |
| 11 | MAL-ED India | 25305300 | India | 2010-2012 | Cohort | 247 | GP | <1 mo | 33.1 (26.1, 35.1) |
| 12 | MISAME I | 18996870 | Burkina Faso | 2004-2006 | RCT | 1,221 | GP | <1 mo | 11.4 (3.3, 12.2) |
| 13 | MISAME II | 19812173 | Burkina Faso | 2006-2008 | RCT | 1,084 | GP | <1 mo | 11.8 (8.2, 12.3) |
| 14 | MISAME III | 36745684 | Burkina Faso | 2019-2020 | RCT | 1,608 | GP | <1 mo | 10.7 (8.0, 11.0) |
| 15 | MRCG Keneba | 26559544;<br>30753251 | Gambia | 2005-2016 | Cohort | 560 | GP | <1 mo | 12.3 (3.8, 19.2) |
| 16 | Pemba | 29163938;<br>34999881 | Tanzania | 2014-2018 | Cohort | 4,109 | GP | <1 mo | 36 (32.5, 37.1) |
| 17 | PM2A<br>Guatemala | 30184223 | Guatemala | 2011-2015 | RCT | 4,212 | GP | <1 mo | 22.9 (22.7, 23.0) |
| 18 | PNS | 17409323;<br>34151973 | Tanzania | 2001-2004 | RCT | 7,956 | GP | <1 mo | 11.6 (10.7, 16.9) |
| 19 | PROMIS<br>Burkina Faso | 31454347 | Burkina Faso | 2015-2017 | RCT | 2,113 | GP | 0-17 mo | 17.7 (17.2, 18.0) |
| 20 | PROMIS Mali | PMC5343313;<br>PMC6711497 | Mali | 2015-2017 | RCT | 1,132 | GP | 6-23.9 mo | 18 (18, 18.1) |
| 21 | ROSE | 34375336 | Niger | 2014-2015 | RCT | 2,507 | GP | 1.4-1.8 mo | 23.8 (23.3, 24.3) |
| 22 | South Africa<br>Study | 37058529 | South Africa | 2018-2021 | RCT | 386 | GP | <1 mo | 25.8 (8.8, 30.2) |
| 23 | ZINC 720 | 27489011 | Burkina Faso | 2010-2012 | RCT | 6,245 | GP | 6-27 mo | 3.6 (0, 7.1) |
| 24 | 3 foods for<br>MAM | 22170366;<br>23256140;<br>25419681 | Malawi | 2009-2011 | RCT | 2,707 | A-S | 6-59 mo | 0.5 (0.5, 1.5) |
| 25 | 10% milk<br>RUTF | 20980648 | Malawi | 2008-2009 | RCT | 1,873 | A-S | 6-59 mo | 1 (0.5, 1.4) |
| 26 | Abx for SAM | 23363496 | Malawi | 2009-2011 | RCT | 2,742 | A-S | 6-59 mo | 1 (0.5, 1.5) |
| 27 | ComPAS | 29690916;<br>32645109;<br>33534818 | Kenya, South Sudan | 2017-2018 | RCT | 4,072 | A-S | 6-59 mo | 2.1 (1.2, 3.2) |
| 28 | DIVIDS | 21628364 | India | 2007-2010 | RCT | 2,079 | A-S | <1 mo | 6 (5.1, 6) |
| 29 | HI MAM | 33963734 | Sierra Leone | 2018-2020 | RCT | 1,286 | A-S | 6-59 mo | 5.5 (5.5, 5.5) |
| 30 | CHAIN | 35427524 | Kenya, Uganda, Malawi,<br>Burkina Faso, Bangladesh,<br>Pakistan | 2016-2019 | Cohort | 3,101 | I-S | 2-23 mo | 6.1 (5.8, 6.4) |
| 31 | SMART 0-6<br>Phase1 | 37182535;<br>26608641 | Uganda | 2018- | Cohort | 2,647 | I-S | 0-6 mo | 6 (6, 6) |
| 32 | SMART 6-60<br>Phase0 | 37182535;<br>26608641 | Uganda | 2017- | Cohort | 1,273 | I-S | 6-60 mo | 6 (6, 6) |
| 33 | SMART 6-60<br>Phase1 | 37182535;<br>26608641 | Uganda | 2018- | Cohort | 3,767 | I-S | 6-60 mo | 6 (6, 6) |

<sup>2</sup> Categorized based on inclusion and exclusion criteria of individual studies. Anthropometry selected (A-S): enrolment based on an anthropometric deficit; Illness selected (I-S): enrolment based on the presence of an illness; General Population (GP): study population enrolled from a general population without consideration of an anthropometric deficit or the presence of illness; other inclusion criteria were however, implemented.

Study populations were categorized into three types: General Population (GP) when enrolment to the original study was not based on an anthropometric deficit or the presence of an illness even if some additional inclusion criteria may have been implemented; and Anthropometry-Selected (A-S) or Illness-Selected (I-S) if the original study enrolled children based on an anthropometric deficit or presence of signs or symptoms of an illness, respectively. For the categorization of geographical regions of study implementation, we used WHO world regions (https://www.who.int/countries).

#### Data cleaning

The pooled dataset underwent a data cleaning process before analysis (Figure 1). Datasets with one row per child (wide format) were re-structured into a long format with one row for each observation at different time-points of a child (and thus multiple rows per child). All duplicate rows were omitted from the dataset (949 observations). Some duplicates occurred because continuous variables in some datasets were recorded with lower precision, e.g., age was received as age in months with one decimal place such that follow-up visits more frequent than 0.1 months could not be differentiated. All observations for individual visits without information on age (22,342 observations) or sex (1,047 observations) were omitted from the analysis. We created a missing category for variables with missing values in individual participants but that were generally captured in the study.

Measurements of exposures that occurred after age 59 months were omitted from the analyses (n = 160); age at death after age 59 months was retained. For 16 children, an age <0 was recorded; these observations were omitted from analyses. Anthropometric measurements were converted to z-scores for length/ height-for-age (HAZ), weight-for-age (WAZ) and weight-for-length/-height (WHZ) according to the WHO Child Growth Standards (27) using the zscore06 command in Stata. Mid-upper arm circumference (MUAC) was included as an absolute measure (in mm). Cut-offs for outliers were decided before commencing the analyses (MUAC >300mm, WAZ >10 and <-10, length <35cm, HAZ >10 and <-12 as well as WHZ >20 and <-10) and individual records were examined; consequently, 54 values (i.e., weight, length, or MUAC) and the corresponding Z-score were re-assigned as ‘Missing’. For dichotomous variables, observations with values other than 0 and 1 were recoded after confirmation with study PI or deleted (2 observations re-assigned as ‘Missing’).

### Variable availability and definition

For each variable (Table 2), a defined format was requested from each study team. In case of deviation, a detailed description of the discrepancy to the requested format, definition and/or assessment method was provided by the study team. The requested format and the availability of data in each study is shown in Supplemental Tables S3 and S4.

**Table 2:**
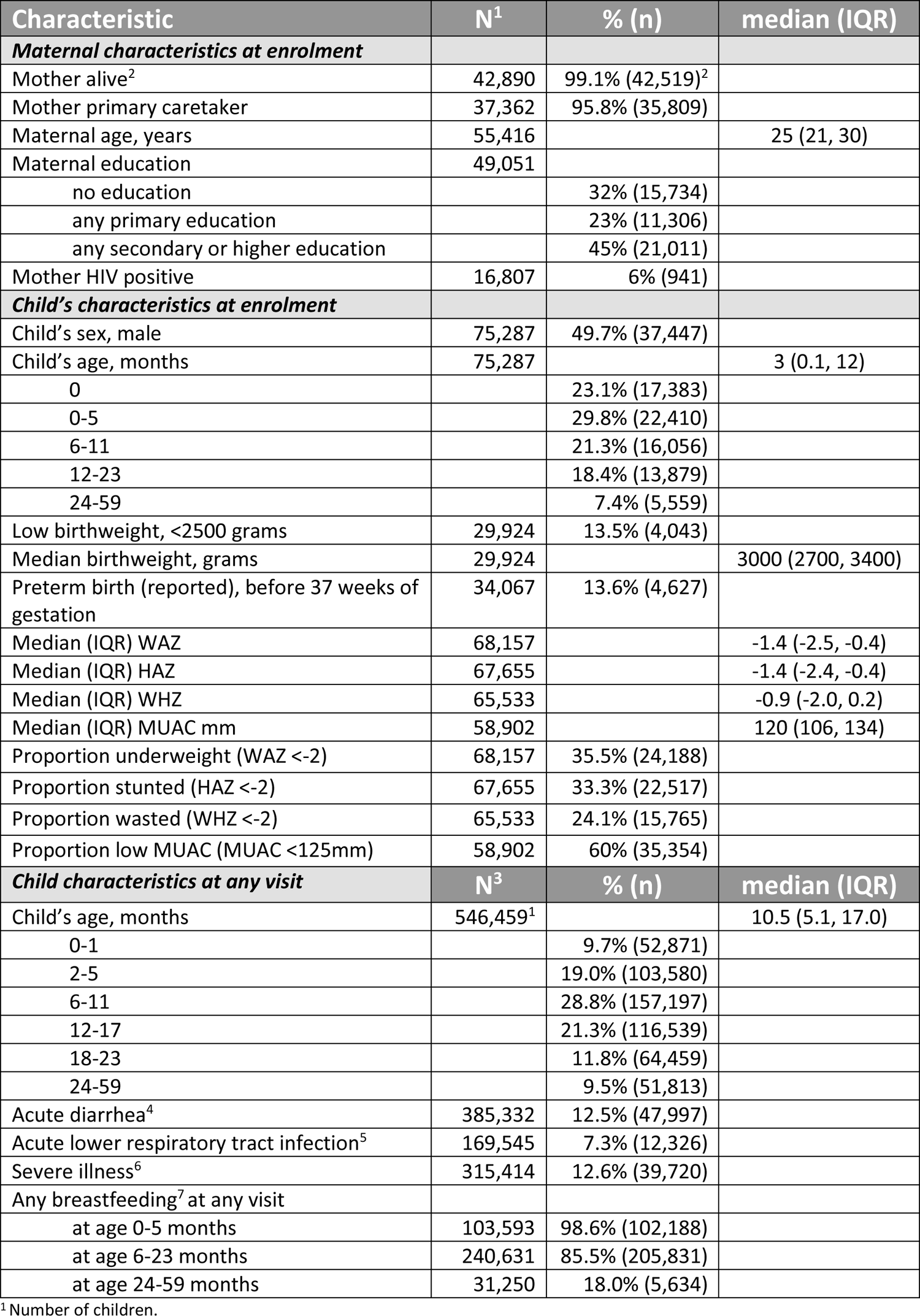
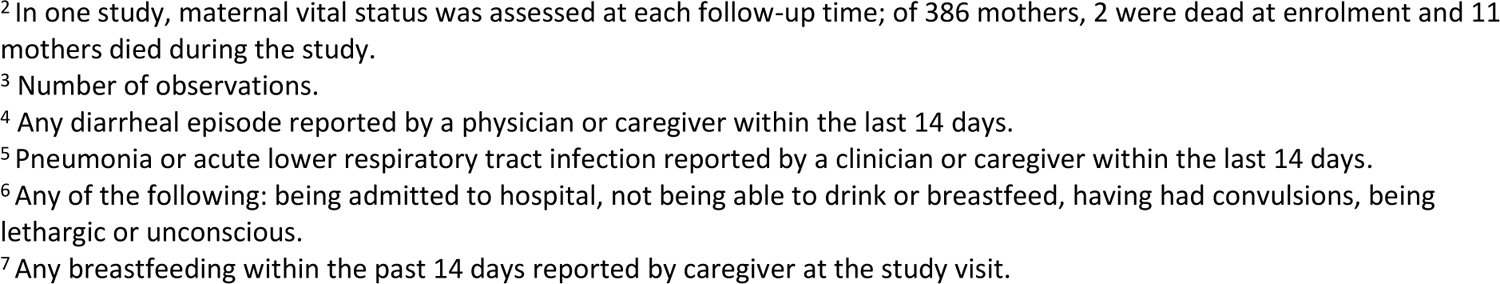
Maternal and child characteristics in the WHO Child Mortality Risk Stratification Multi-Country Pooled Cohort (WHO-CMRS)

| Characteristic | N<sup>1</sup> | % (n) | median (IQR) |
| --- | --- | --- | --- |
| **Maternal characteristics at enrolment** |  |  |  |
| Mother alive<sup>2</sup> | 42,890 | 99.1% (42,519)<sup>2</sup> |  |
| Mother primary caretaker | 37,362 | 95.8% (35,809) |  |
| Maternal age, years | 55,416 |  | 25 (21, 30) |
| Maternal education | 49,051 |  |  |
| no education |  | 32% (15,734) |  |
| any primary education |  | 23% (11,306) |  |
| any secondary or higher education |  | 45% (21,011) |  |
| Mother HIV positive | 16,807 | 6% (941) |  |
| **Child's characteristics at enrolment** |  |  |  |
| Child's sex, male | 75,287 | 49.7% (37,447) |  |
| Child's age, months | 75,287 |  | 3 (0.1, 12) |
| 0 |  | 23.1% (17,383) |  |
| 0-5 |  | 29.8% (22,410) |  |
| 6-11 |  | 21.3% (16,056) |  |
| 12-23 |  | 18.4% (13,879) |  |
| 24-59 |  | 7.4% (5,559) |  |
| Low birthweight, <2500 grams | 29,924 | 13.5% (4,043) |  |
| Median birthweight, grams | 29,924 |  | 3000 (2700, 3400) |
| Preterm birth (reported), before 37 weeks of gestation | 34,067 | 13.6% (4,627) |  |
| Median (IQR) WAZ | 68,157 |  | -1.4 (-2.5, -0.4) |
| Median (IQR) HAZ | 67,655 |  | -1.4 (-2.4, -0.4) |
| Median (IQR) WHZ | 65,533 |  | -0.9 (-2.0, 0.2) |
| Median (IQR) MUAC mm | 58,902 |  | 120 (106, 134) |
| Proportion underweight (WAZ <-2) | 68,157 | 35.5% (24,188) |  |
| Proportion stunted (HAZ <-2) | 67,655 | 33.3% (22,517) |  |
| Proportion wasted (WHZ <-2) | 65,533 | 24.1% (15,765) |  |
| Proportion low MUAC (MUAC <125mm) | 58,902 | 60% (35,354) |  |
| **Child characteristics at any visit** |  |  |  |
| Child's age, months | 546,459<sup>1</sup> |  | 10.5 (5.1, 17.0) |
| 0-1 |  | 9.7% (52,871) |  |
| 2-5 |  | 19.0% (103,580) |  |
| 6-11 |  | 28.8% (157,197) |  |
| 12-17 |  | 21.3% (116,539) |  |
| 18-23 |  | 11.8% (64,459) |  |
| 24-59 |  | 9.5% (51,813) |  |
| Acute diarrhea<sup>4</sup> | 385,332 | 12.5% (47,997) |  |
| Acute lower respiratory tract infection<sup>5</sup> | 169,545 | 7.3% (12,326) |  |
| Severe illness<sup>6</sup> | 315,414 | 12.6% (39,720) |  |
| Any breastfeeding<sup>7</sup> at any visit |  |  |  |
| at age 0-5 months | 103,593 | 98.6% (102,188) |  |
| at age 6-23 months | 240,631 | 85.5% (205,831) |  |
| at age 24-59 months | 31,250 | 18.0% (5,634) |  |

<sup>1</sup> Number of children.
<sup>2</sup> In one study, maternal vital status was assessed at each follow-up time; of 386 mothers, 2 were dead at enrolment and 11 mothers died during the study.
<sup>3</sup> Number of observations.
<sup>4</sup> Any diarrheal episode reported by a physician or caregiver within the last 14 days.
<sup>5</sup> Pneumonia or acute lower respiratory tract infection reported by a clinician or caregiver within the last 14 days.
<sup>6</sup> Any of the following: being admitted to hospital, not being able to drink or breastfeed, having had convulsions, being lethargic or unconscious.
<sup>7</sup> Any breastfeeding within the past 14 days reported by caregiver at the study visit.

#### Anthropometry

Moderate anthropometric deficit is defined as a Z-score <-2 and ≥-3, and a severe deficit as a Z-score <-3. MUAC is included as a simple measurement (mm) and categorized as ≥125 mm (no deficit), 115-124 mm (moderate deficit) and <115 mm (severe deficit) (28). For children 0-1 month of age, we also applied cut-offs at 115 mm and 105 mm, and for children 2-5 months, 120mm and 110mm, respectively. Other anthropometric measures such as height-for-age difference (HAD in cm) as well as changes in HAD (29), growth velocity (30), and indicators suggested in NICE guidelines (31) were estimated.

#### Morbidity

All morbidity variables were dichotomous (yes/no). Haemoglobin concentration (g/dL), peripheral arterial oxygen saturation (%), and blood glucose concentration (mmol/L) were included as continuous variables. The main morbidity variables used in the primary analyses were diarrhoea, acute lower respiratory tract infection (ALRTI) and severe illness. Twenty-nine studies had information on diarrhoea. In 3 studies, diarrhoea was assessed by a physician or trained field worker; in the other 24 studies information was from parental report of the child having any episode of diarrhoea within the last 14 days. Study definitions of LRTI were used: in 5 studies, LRTI referred to children diagnosed with pneumonia by a physician; in 10 studies, this referred to children reported by caretakers to have pneumonia or acute lower respiratory tract infection according to WHO definitions based on symptoms in the last 14 days (WHO classification of signs indicating pneumonia have been revised between 2000 and 2023; studies applied classifications recommended at the time of study implementation). The WHO definitions of pneumonia are sensitive and not specific and include fast breathing or lower chest indrawing and a history of difficulty breathing (32–34). Severe illness was defined as being admitted to hospital or having any of the WHO IMCI danger signs, i.e., the child is not able to drink or breastfeed, the child has had convulsions, and the child is lethargic or unconscious (34). The IMCI danger sign of ‘vomiting everything’ was not available in any of the studies. Fifteen datasets had data to generate this dichotomous severe illness variable. In addition, we created a variable that tallied the total number of severe illness characteristics that were present (range 0 to 4). Other morbidity variables were available (see Table 2) but not used in primary analyses as they were setting-specific, only available in a minority of children in the pooled cohort (<50%), anticipated to have a low sensitivity and/or specificity for mortality, or of secondary interest.

#### Other risk exposures

Data on “any breastfeeding” practices was available from 26 studies; in 16 studies was defined as any breastfeeding within the past 24 hours reported by the caregiver. Two studies reported breastfeeding practices over longer intervals, i.e., the past 3-4 days (study no. 5) and the past month (study no. 4). Descriptions of this variable were not detailed in the remaining eight studies. The information if the child was exclusively breastfed (i.e. no other foods or fluids than breastmilk was given) was available in 16 studies. The following non-time-varying variables were also available for inclusion in the primary analyses: low birthweight (LBW; birthweight <2500g; 17 studies), pre-term birth (PTB; <37 completed weeks of gestation at birth; 13 studies), mother as primary caregiver (14 studies), maternal age (25 studies), maternal educational level at enrolment (20 studies, respectively), and rural vs. urban residence (33 studies). Methods used to establish gestational age varied by study and included date of last menstrual period, early ultrasound, and study-specific algorithms. We categorized maternal educational level as no, primary, and secondary education and above. In case of continuous variables, we used the cut-offs of 0, 1-6 and >6 years of education to categorize the level accordingly.

### Principles and plans for statistical analysis

The RSWG agreed that future analyses will use all-cause mortality as the main outcome, accounting for repeated measurements in the individual child, clustering within studies, differences in age at inclusion, as well as duration and frequency of follow-up in the individual studies. It was also agreed that all analyses should be presented by age group and sex. RCTs are considered as single parallel cohorts with no adjustment for effect/ no effect of intervention. As per definition, the interventions in each study were randomly assigned independent of the exposures in our analyses. Any intervention was therefore considered equally to any co-intervention.

The aim is to report absolute risk and risk differences (as percentage or percentage points) with 95% confidence intervals in addition to relative measures, such as relative risk or odds ratio. Widely used tools to improve the quality of reporting, such as the “Strengthening the Reporting of Observational Studies in Epidemiology” (STROBE) statement, recommend that both relative and absolute measures of association together with their level of precision are reported. The rationale for this is that relative measures alone can be misleading due to differences in baseline risk (risk in the reference group) obscuring the clinical significance of the effect; absolute risks can therefore be highly informative for decision-making (35).

Age- and sex-specific mortality risk for each selected exposure will be estimated individually. An interaction term between the exposure and age will be included in the statistical models because we assumed that the association between the exposures and death are modified by age. Results will be presented by the following age groups: 0-6, 6-11, 12-24, and 24-59 months. When data allows, and especially for specific analyses focusing on children under 6 months of age, age groups will be further refined. Further, the interaction of each anthropometric predictor with other predictors will be measured on an additive scale. We will also examine the extent to which study-level characteristics, such as study population type and geography modify the association between selected predictors and the risk of death.

Not all studies have data on all possible risk exposures. To understand how individual studies potentially drive estimates, forest plots will be used to visually represent the data characteristics of studies. Sensitivity analyses will be performed to understand the effect of outliers or missingness. For each analysis, a detailed statistical analysis plan will be developed and reported with findings.

### COHORT CHARACTERISTICS

The pooled dataset includes 75,287 children (49.7% male) with 546,459 observations between 0 and 59 months of age. Twenty-nine percent of all observations were between birth and 5 months of age (156,451 observations), 29% at ages 6-11 months (157,197 observations), 33% at age 12-23 months (180,998 observations), and 9% at age 24-59 months (51,813 observations). Children from sub-Saharan Africa make up the largest part of the pooled cohort data (n = 58,581; 78%). Studies in South-East Asia included 8,756 children (12%), and 7,952 children were from other world regions (11%).

Two thirds (66%, n= 49,740) of all children were enrolled into 23 studies that did not use a health condition or anthropometry as an inclusion criterion (GP), 20% (n= 14,759) were enrolled in six studies based on an anthropometric deficit (A-S), and 14% (n= 10,788) in four studies based on the presence of an illness (I-S). Sixteen (48%) of the included studies were birth cohorts (15 of the GP, one of the A-S and none of the I-S studies) with a total of 32,284 children. Of the 16 studies defined as birth cohorts, 12 studies enrolled pregnant women, one study enrolled neonates within 12 hours and 1 study within 48 hours after birth (study nos. 4 and 28; Table 1). For two studies, the study protocol allowed for enrolment up to 17 days after birth (nos. 10 and 11; Table 1). The median length of follow-up of the individual children was 15 months (IQR 7.6 to 18.4 months); follow-up time was longer in the GP studies (median 17 months) compared to the A-S and I-S populations (3 and 6 months, respectively). The number of scheduled visits varied substantially between studies, ranging from 2 to 81, with a median of 12 visits (IQR 7-19) in the pooled cohort. The median number of visits differed between study population types: 14 in the GP, 6 in the A-S and 4 in the I-S studies.

Table 2 summarizes characteristics of the children and their mothers included in the pooled cohort. The median age at enrolment was 3 months; 23% (n = 17,383) children were enrolled within 17 days of birth, and 49% (n =36,648) children before the age of two months. GP studies enrolled children at younger ages; the median age at enrolment was 1 month in the GP studies, 13 months in the A-S studies, and 11 months in the I-S studies.

At enrolment, 33% of children were stunted (HAZ <-2), 24% were wasted by WHZ (WHZ <-2), 60% were wasted by MUAC (MUAC <125mm), and 36% underweight (WAZ <-2). The anthropometric status differed considerably between included studies and age-groups both at enrolment (Tables S5, S6a, S6b) and throughout the study period (Figure S1). However, due to the heterogeneity in inclusion criteria as well as ages at enrolment of the individual studies, it should be noted that the means in each of these study population or age-groups depend on the varying contributions from individual studies.

Twelve percent of all children were reported to have an episode of diarrhoea within the prior 14 days (at any visit). When groups were analysed based on study population type, the prevalence of diarrhoea was somewhat lower for the youngest age group (<6 months) and then relatively stable at around 7% (GP children), 32% (A-S children) and 50% (I-S children), respectively. For LRTI, the prevalence in GP children varied between 5% for children younger than 12 months and 1% for older children. Among studies categorized as I-S, the prevalence was relatively stable at around 40%. LRTI was not captured in any of the A-S studies.

The proportion of children with low birthweight was 13% in GP, 5% in A-S and 21% in I-S studies. However, only one study in each of the A-S (1,112 children) and I-S cohorts (2,145 children) had information on birthweight. None of the A-S studies reported data on gestational age at birth. In GP studies, the prevalence of reported preterm birth was 16%, and in the I-S studies it was 4%. The proportion of children currently receiving any breastfeeding reduced from almost universal practice in the age group <6 months (91-99%) to less than 20% in the oldest age group (24-59 months).

Children enrolled in studies with inclusion criteria based on having an illness (I-S) were less likely to be breastfed at any age (Figure 2). Overall, mothers had a median age of 24 years (IQR 20-28.5 years); 33% had no formal education, 20% had primary education and 47% had an educational level of secondary school or above.

**Figure 2:**
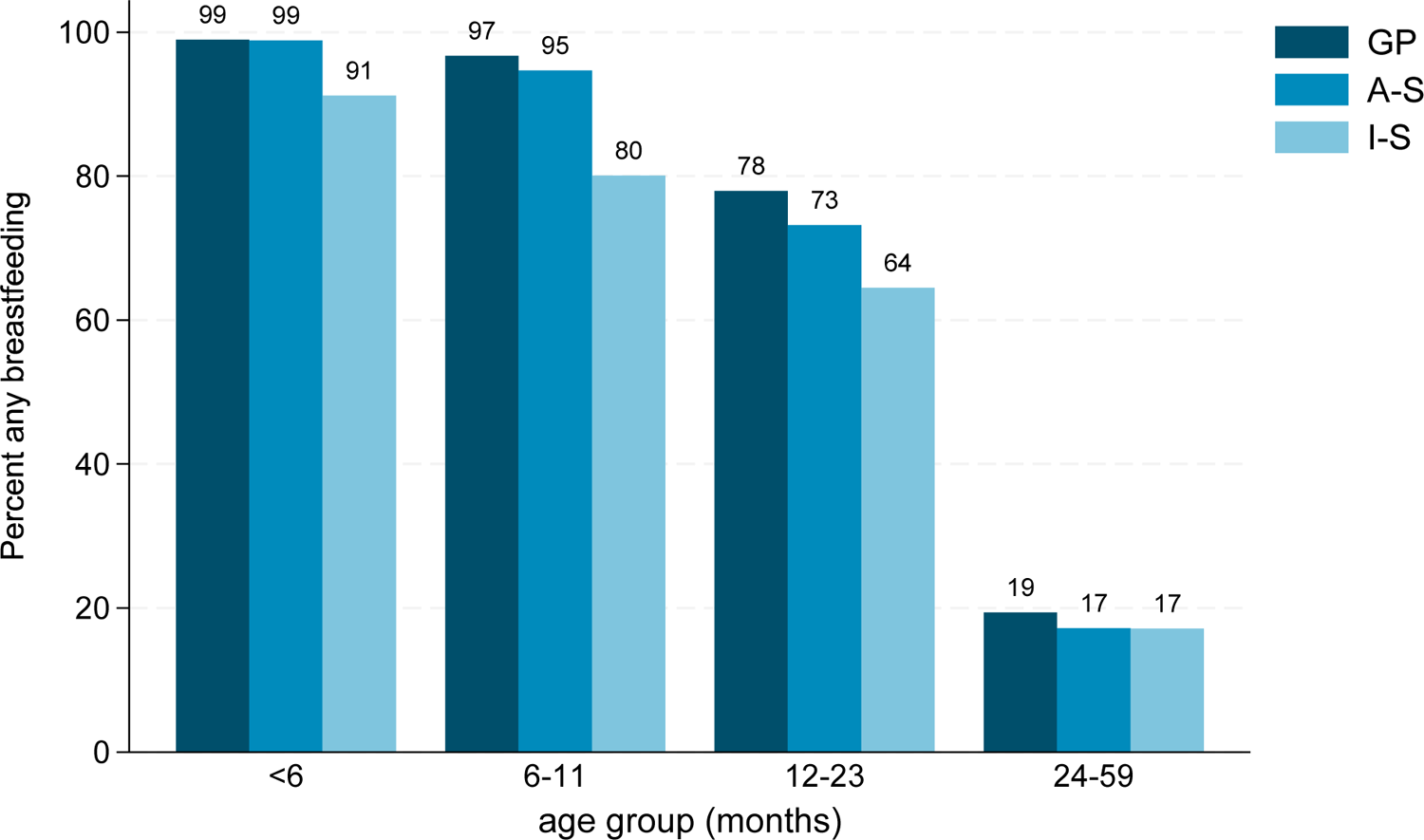
Proportion of children in the WHO Child Mortality Risk Stratification Multi-Country Pooled Cohort (WRS-CMRS) being breastfed at time of follow-up according to age group and study population type; GP (not enrolled by anthropometric deficit or illness), A-S (anthropometry-selected), and I-S (illness-selected)

There were 2,805 children who died within the study period. The proportion of children who died differed according to how studies enrolled their population: 2.5% (n = 1,264) in the GP studies, 2.5% (n = 366) in the A-S studies, and 11.1% (n = 1,201) in the I-S studies. There were 145 children (all from GP studies) for whom the age at death was not documented. For these children, we estimated the age at death by adding half of the median follow-up time for the original study to the age at last measurement. The distribution of deaths by age (frequency) and by study population type is presented in Figure 3. (Note: Age distribution of deaths also reflects age at enrolment). The median observed follow-up time from enrolment to death was 1 month (IQR 0.2, 4.7 months) for the pooled cohort; with 4.1 (0.8, 9.5) months in the GP studies, 1 (0.5, 2.3) month in the A-S studies and 0.3 (0.1, 1.4) months in the I-S studies. Among children with available information on HIV status (16,190 children), 653 were reported to be HIV infected (4%); of these 653 children, 140 (21%) died during the study period.

**Figure 3:**
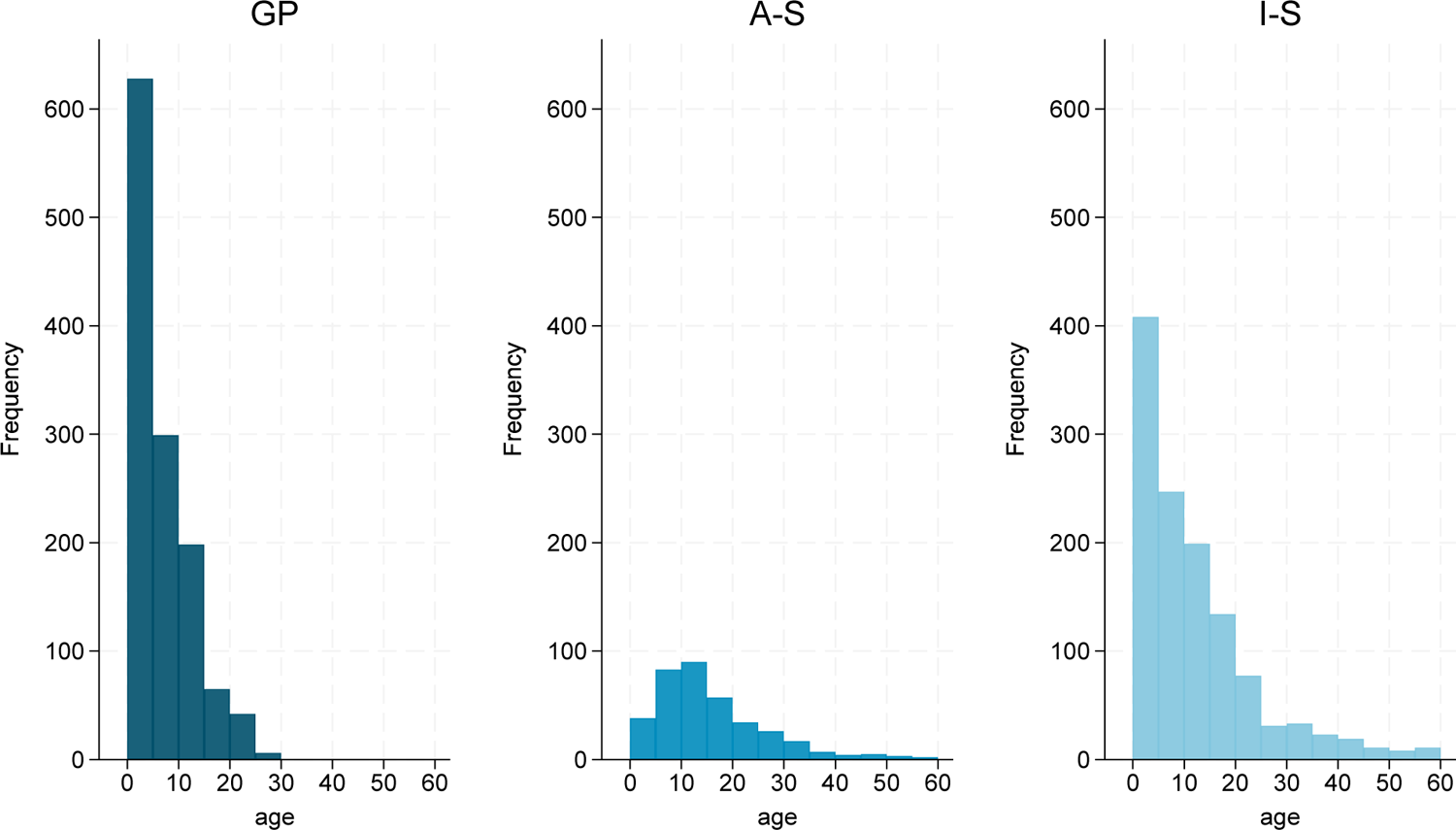
Distribution deaths in the WHO Child Mortality Risk Stratification Multi-Country Pooled Cohort (WHO-CMRS) by age (months) and study population type

## CONCLUDING REMARKS

The aim of this cohort profile is to describe the process of assembling the WHO-CMRS cohort as well as the characteristics of included studies and enrolled children. As expected, children enrolled to studies based on anthropometric criteria or the presence of illness differed from those resembling the general population. In addition to anthropometry and morbidity status, there were differences in individual characteristics such as age at enrolment and study design features such as number and frequency of scheduled study visits or assessment methods such as determining gestational age.

These differences will need to be taken into account in future analyses. Although the heterogeneity of included studies posed some challenges in the pooling of datasets and interpretation of findings, it also provides opportunities to explore specific differences these characteristics through sensitivity analyses.

## FURTHER DETAILS

### Data sharing statement

All contributing research teams have acknowledged that the pooled data can only be used for collaborative activities within the WHO RSWG, with no transfer of ownership. Respective Principal Investigators (see Supplementary Table S1) can be contacted for access to their data.

### Funding declaration

WHO gratefully acknowledges funding support from USAID to establish the pooled database and conduct the main analyses.

### Contributors statement

– Conceptualization: Rajiv Bahl, Nigel Rollins
– Study design: Alemayehu Argaw, Rajiv Bahl, James A. Berkley, Ranadip Chowdhury, Siri Kaldenbach, Nigel Rollins, Catherine Schwinger, Tor Strand, Judd Walson with input from all members of the WHO-RSWG
– Data cleaning and preparation: Siri Kaldenbach, Catherine Schwinger, Tor Strand
– Statistical analysis: Catherine Schwinger, Tor Strand
– Data acquisition and interpretation: all
– Drafting the manuscript: Catherine Schwinger, Tor Strand, Siri Kaldenbach, Nigel Rollins, Jay Berkley
– Reviewing the manuscript: all

### Disclaimer

The authors alone are responsible for the views expressed in this paper and they do not necessarily represent the views, decisions, or policies of the institutions with which they are affiliated.

## Data Availability

Principle Investigators of the individual studies can be contacted for a request to access their data.

## Notes

### Competing Interest Statement

The authors have declared no competing interest.

### Author Declarations

The WHO Ethics Review Committee granted an exemption in March 2022 with protocol ID: ERC.0003745

