## Supplementary Material for "Cohort profile: The WHO Child Mortality Risk Stratification Multi-Country Pooled Cohort (WHO-CMRS) to identify predictors of mortality through early childhood"

**Table S1**: List of participants in the Risk Stratification Working Group (RSWG)*

| | Name of person | Institution/Affiliation |
|---|---|---|
| 1 | Ahmed, Tahmeed | International Centre for Diarrhoeal Disease Research, Bangladesh |
| 2 | Argaw, Alemayehu | Ghent University |
| 3 | Bahl, Rajiv | World Health Organization |
| 4 | Bailey, Jeanette | International Rescue Committee (IRC) |
| 5 | Baqui, Abdullah | Johns Hopkins University |
| 6 | Becquey, Elodie | International Food Policy Research Institute |
| 7 | Berkley, Jay | Kenya Medical Research Institute (KEMRI) / Wellcome Trust Research Programme, Kilifi, Kenya |
| 8 | Brown, Kenneth | University of California, Berkley |
| 9 | Chisti, Mohammed Jobayer | International Centre for Diarrhoeal Disease Research, Bangladesh |
| 10 | Chowdhury, Ranadip | Society for Applied Studies, New Delhi |
| 11 | Diallo, Hama A. | University Joseph KI-ZERBO, Burkina Faso |
| 12 | Duggan, Christopher | Harvard T.H. Chan School of Public Health and Boston Children's Hospital |
| 13 | Evans, Denise | University of the Witwatersrand, Johannesburg, South Africa |
| 14 | Fawzi, Wafaie | Harvard T.H. Chan School of Public Health |
| 15 | Fazal, Ali | International Centre for Diarrhoeal Disease Research, Bangladesh |
| 16 | Goga, Ameena | World Health Organization |
| 17 | Guindo, Ousmane | Epicentre, MSF |
| 18 | Grais, Rebecca | Pasteur Institute, Paris, France |
| 19 | Hamer, David | Boston University |
| 20 | Hess, Sonja | University of California, Davis |
| 21 | Huybregts, Lieven | International Food Policy Research Institute |
| 22 | Isanaka, Sheila | Harvard T.H. Chan School of Public Health and Epicentre |
| 23 | Jehan, Fyezah | Aga Khan University |
| 24 | Kabakyenga, Jerome | Mbarara University of Science and Technology, Mbarara, Uganda |
| 25 | Kabore, Patrick | World Health Organization |
| 26 | Kaldenbach, Siri | Innlandet Hospital Trust |
| 27 | Kerac, Marko | London School of Hygiene and Tropical Medicine |
| 28 | Khanam, Rasheda | Johns Hopkins University |
| 29 | Kissoon, Niranjan | University of British Columbia |
| 30 | Kounnavong, Sengchanh | Institute for Tropical and Public Health Institute, Vientiane, Lao PDR |
| 31 | Lachat, Carl | Ghent University |
| 32 | LaGrone, Lacey | University of Colorado |
| 33 | Le Port, Agnes | French National Research Institute for Sustainable Development |
| 34 | Lelijveld, Natasha | Emergency Nutrition Network |
| 35 | Leroy, Jef | International Food Policy Research Institute |
| 36 | Mahfuz, Mustafa | International Centre for Diarrhoeal Disease Research, Bangladesh |
| 37 | Maleta, Kenneth | Kamuzu University of Health Sciences |
| 38 | Manary, Mark | Washington University in St. Louis |

| | | |
|---|---|---|
| 39 | Manji, Karim P. | Muhimbili University of Health and Allied Sciences |
| 40 | Marconi, Sam | Christian Medical College Vellore |
| 41 | McGrath, Marie | Emergency Nutrition Network |
| 42 | Mohan, Venkata Raghava | Christian Medical College Vellore |
| 43 | Moore, Sophie | King's College London |
| 44 | Mugisha, Nathan Kenya | Walimu, Kampala, Uganda |
| 45 | Mupere, Ezekiel | Makarere University School of Medicine, Uganda |
| 46 | Mwaringa, Shalton | Kenya Medical Research Institute (KEMRI) / Wellcome Trust Research Programme, Kilifi, Kenya |
| 47 | Nisar, Yasir Bin | World Health Organization |
| 48 | Natarajan, Sindhu Kulandaipalayam | Christian Medical College Vellore, India |
| 49 | Ngari, Moses | Kenya Medical Research Institute (KEMRI) / Wellcome Trust Research Programme, Kilifi, Kenya |
| 50 | Nisar, Imran | Aga Khan University, Pakistan |
| 51 | Olney, Deanna | International Food Policy Research Institute |
| 52 | Ouedraogo, Jean-Bosco | Health Sciences Research Institute, Bobo-Dioulasso, Burkina Faso |
| 53 | Prentice, Andrew | MRC Unit, The Gambia at London School of Hygiene and Tropical Medicine, Banjul, The Gambia |
| 54 | Prost, Audrey | University College London |
| 55 | Roberfroid, Dominique | Belgian Health Care Knowledge Centre (KCE), Brussels |
| 56 | Rockers, Peter | Boston University |
| 57 | Rollins, Nigel | World Health Organization |
| 58 | Ruel, Marie | International Food Policy Research Institute |
| 59 | Saleem, Ali | Aga Khan University, Pakistan |
| 60 | Sazawal, Sunil | Johns Hopkins University |
| 61 | Schwinger, Catherine | University of Bergen |
| 62 | Singa, Benson | Kenya Medical Research Institute (KEMRI) / Wellcome Trust Research Programme, Kilifi, Kenya, and University of Washington |
| 63 | Stobaugh, Heather | International Rescue Committee and Action Against Hunger |
| 64 | Strand, Tor A. | Innlandet Hospital Trust, University of Bergen |
| 65 | Taneja, Sunita | Society for Applied Studies |
| 66 | Timbwa, Molly | Kenya Medical Research Institute (KEMRI) / Wellcome Trust Research Programme, Kilifi, Kenya |
| 67 | Toe, Laéticia Céline | University of Gent |
| 68 | Trehan, Indi | University of Washington |
| 69 | Trilok-Kumar, Geeta | Delhi University and Ashoka University |
| 70 | Voskuijl, Wieger | Emma Children's Hospital, Amsterdam UMC, University of Malawi |
| 71 | Walson, Judd | Johns Hopkins University |
| 72 | Wang, Donqing | Harvard T.H. Chan School of Public Health and George Mason University |
| 73 | Waseem, Arup Ali | Center for Public Health Kinetics, New Delhi, India |
| 74 | Wiens, Matthew | University of British Columbia |

*The RSWG includes researchers with responsibility for the primary data (in general 2 persons per dataset) plus persons who joined the working group meetings and made substantive technical input to the RS concepts or pooled analysis approach.

**Table S2: Description of studies included in the WHO Child Mortality Risk Stratification Multi-Country Pooled Cohort (WHO CMRS)**

| Nr | Short title | Study site | Urban/Rural | Classification and description of inclusion criteria | Type of intervention (*category*[1] and description) | Primary study outcome | Total follow-up OR follow up frequency |
|---|---|---|---|---|---|---|---|
| 1 | AMANHI Bangladesh | Health facilities in Sylhet area, Bangladesh | | GP: Pregnant women (8-19 weeks of gestation) | N/A | Uncover biological markers as predictors of important maternal and foetal outcomes | Up to 42days postpartum |
| 2 | AMANHI Pakistan | Health facilities in Karachi area, Pakistan | | GP: Pregnant women (8-19 weeks of gestation) | N/A | Uncover biological markers as predictors of important maternal and foetal outcomes | Up to 42d postpartum |
| 3 | CARING | Villages in catchment area, India | Rural | GP: Pregnant women with their children living in study area. | Community mobilization through participatory women's groups<br><br>Health and nutrition counselling through home visits | Children's mean LAZ at 18 months | Up to 24m postpartum |
| 4 | CHILD 2 | Clinics and Hospital, Tanzania | Peri-urban | GP: Pregnant women <34w, or from labour ward. Singleton baby. | *Nutritional:*<br>(1) zinc, (2) multivitamins, (3) multivitamin + zinc, (4) placebo<br><br>All: standard care | Incidence of clinical symptoms of diarrhoea and lower respiratory infection | Monthly, total 18 months. |
| 5 | CRI study | Clinics in study area, India | Urban | GP: Recruited at birth from women living in area (slum) identified by household survey who intended to stay in area for 3 years. | N/A | Overall rotavirus infections | Every 2 weeks, up to 26 months |
| 6 | IECDZ (Improving Early Childhood | Villages in Zambia | Rural | GP: Household in randomly selected villages with child aged | *Caregiver support:*<br>Intervention clusters: 2 visits/month during 1st study | Children's stunting (HAZ <-2) and neurocognitive development | After 1 year and 2 years. |

| | | | | | | | |
|---|---|---|---|---|---|---|---|
| | Development in Zambia) study | | | 6-12 m at baseline, female caregivers (>15y). | year by child development agents (CDAs); invited to fortnightly parenting group meetings (led by trained "head-mothers" from communities) | | |
| 7 | iLiNS | Health facilities in Dandé Health District, Burkina Faso | Rural | GP: Community screening: 9-month-old infants, living in area, planned to stay. | *Nutritional:*<br>(1) SQ-LNS without zinc, and placebo tablet, (2) SQ-LNS with 5 mg zinc, and placebo tablet, (3) SQ-LNS with 10 mg zinc, and placebo tablet, (4) SQ-LNS without zinc, and 5 mg zinc tablet<br>(5) no intervention | Weekly morbidity assessment for incidence of diarrhoea and malaria; growth and plasma zinc concentration | 9 months (from age 9 months to 18 months) |
| 8 | Lao zinc | Health facilities in Khammouane Province, Lao PDR | Rural | GP: Community screening: Households with potentially eligible children (6-23m) were screened in study area and accept home visits. | *Nutritional:*<br>(1) daily preventive zinc tablet;<br>(2) daily preventive multiple micronutrient powder;<br>(3) therapeutic zinc tablet for diarrhoea;<br>(4) placebo control | Weekly morbidity assessment for diarrhoea incidence; growth (length and weight), diarrhoea incidence, haemoglobin and micronutrient status, innate and adaptive immune function. | Every 16-20 weeks, 9 months total. |
| 9 | Low BW | Health facilities, Burkina Faso | Rural | GP: Pregnant women attending ANC in 3rd trimester. | N/A | Death within 12 months from date of birth | 1 year total. |
| 10 | MAL-ED Bangladesh | Clinics in Bangladesh | Urban | GP: Community screening: Enrolled within 17 days after birth, plans stay in study area. | N/A | Overall MAL-ED: Physical growth, cognitive development and immune response to oral and parenteral vaccines. | Every 2 weeks, up to 24 months. |
| 11 | MAL-ED India | Clinics, India | Urban | GP: New-born infants with parent/primary caregiver | N/A | Overall MAL-ED: Physical growth, cognitive development and | Every 2 weeks, up to 24 months. |

| | | | | | | | |
|---|---|---|---|---|---|---|---|
| | | | | being a permanent resident of the study area and those willing to permit home visits. | | immune response to oral and parenteral vaccines. | |
| 12 | MISAME I | Health facilities, Burkina Faso | Rural | GP: Pregnant women living in study area. | *Nutritional (and medical):*<br>(1) Iron and folic acid supplement<br>(2) UNIM-MAP (multiple micronutrient) daily until 3 months after delivery<br><br>Additional randomization to<br>(1) 300mg chloroquine/week<br>(2) 1500mg sulfadoxine + 75mg pyrimethamine once in 2nd and 3rd trimester<br><br>All: standard treatment | Gestational duration, birthweight, birth length and Rohrer's ponderal index (weight/length$^3$) | Monthly visits, 12 months total. |
| 13 | MISAME II | Health facilities, Burkina Faso | Rural | GP: Pregnant women living in study area. | *Nutritional (and medical):*<br>(1) Daily prenatal fortified food supplement (~372 kcal) + MMN, (2) only MMN<br><br>Additional randomization:<br>(1) double dose sulfadoxine-pyrimethamine in 2nd and 3rd trimester<br>(2) triple dose<br><br>All: Standard treatment | Birthweight, birth length and Rohrer's ponderal index (weight/length$^3$) | Monthly |
| 14 | MISAME III | Health facilities, Burkina Faso | Rural | GP: Pregnant women living in study area. | *Nutritional:*<br>2x2 factorial design:<br>(1) prenatal BEP + postnatal BEP<br>(2) prenatal BEP + no postnatal BEP | Prevalence of small for gestational age and HAZ at age 6 months. | Monthly |

| | | | | | | | |
|---|---|---|---|---|---|---|---|
| | | | | | (3) no prenatal BEP + postnatal BEP<br>(4) no intervention<br><br>All: standard IFA tablets | | |
| 15 | MRCG Keneba | Clinics, The Gambia | Rural | GP: Pregnant women and children living in three core villages. | N/A | Growth development, pregnancy, and lactation | 2 monthly |
| 16 | Pemba | Health facilities in Pemba Island, Tanzania | Rural | GP: Pregnant women (8-19 weeks of gestation) | N/A | Uncover biological markers as predictors of important maternal and foetal outcomes. | Up to 42d postpartum |
| 17 | PM2A | Living in study area, Guatemala | | GP: Pregnant women and planning to reside in the area until child reached 24 months of age. | *Nutritional and social:*<br>(1) full FR + CSB; (2) half FR + CSB; (3) no FR + CSB; (4) full FR + SQ-LNS; (5) full FR + MNP; (6) no intervention | Linear growth (length for age z-score and length for age difference). | 1, 4, 6, 9, 12, 18 months. Total 24 months of age. |
| 18 | PNS | Clinics, Tanzania | Urban | GP: Pregnant women attending clinic, negative HIV test, plans to stay in the area until 1 year after delivery, gestational age 12-27 weeks at enrolment | *Nutritional:*<br>(1) multivitamin supplementation, (2) placebo<br><br>All: prenatal supplementation iron and folic acid | Low birth weight (<2500g), preterm delivery (<37w gestational) and foetal death | Every 1 month. Total 18 months. |
| 19 | PROMIS Burkina Faso | Health facilities in Gourcy health district, Burkina Faso | Rural | GP: Singleton child 0-6 weeks of age. | *Nutritional and social:*<br>integrated into existing facility-based program for acute malnutrition screening and referral:<br>(1) Monthly provision of 30 20g sachets of SQ-LNS for daily use) and monthly information on appropriate use; enhanced counselling on child nutrition, hygiene and health | Acute malnutrition screening coverage and incidence. | Monthly, 18 months total. |

| | | | | | | | |
|---|---|---|---|---|---|---|---|
| | | | | | (2) no intervention<br><br>All:<br>technical/financial support for quality implementation of consultations at health centres, quarterly AM screening in community, mobilization of key community members. | | |
| 20 | PROMIS Mali | Health facilities in Bla and San health districts, Mali | Rural | GP: Singleton child 6.0-6.9 months of age. | *Nutritional and social:*<br>integrated into village-based platform for acute malnutrition screening and referral:<br>(1) Monthly provision of 30 20g sachets of SQ-LNSs and monthly information on appropriate use;<br>(2) no intervention<br><br>All:<br>monthly screening meetings, referral of MAM and SAM cases, monthly parental group meetings on behaviour change communication on nutrition, health, and hygiene practices, CHW monthly stipend for monthly meeting. | Acute malnutrition screening coverage and prevalence. | Anthropometry at 9, 12 and 16 months, 18 months total. |
| 21 | ROSE | Hospital, health facilities, Niger | Rural | GP: Community screening of all infants 6-8 weeks of age. | *Medical:*<br>(1) Three doses of BRV-PV (against rotavirus)<br>(2) placebo | Efficacy of three doses of BRV-PV vs. placebo against a first episode of severe rotavirus gastroenteritis. | Up to 2 years of age. |

| | | | | | | | |
|---|---|---|---|---|---|---|---|
| 22 | South Africa Study | Health facilities in catchment area in rural South Africa | Rural | GP: Mother > 18y, child born after 15.12.2017, staying in catchment area. | *Caregiver support:*<br>Support on child health, nutrition, developmental milestones, and appropriate play-based activities by trained Community health workers | Children's stunting (HAZ <-2) and neurocognitive development | Up to 36 months. |
| 23 | ZINC 720 | Health facilities in Orodara health district Burkina Faso | Rural | GP: Community screening: 6-27 months, living in area and no zinc supplementation. | *Nutritional:*<br>(1) 10 mg Zn/day for 10 days every 16 weeks<br>(2) 7 mg Zn/day)<br>(3) 20 mg Zn/day for 10 days for diarrhoea<br>(4) morbidity surveillance control<br>(5) non-intervention control<br><br>All except (5): Weekley morbidity surveillance with treatment of uncomplicated diarrhoea, fever and malaria and referral for other illness. | Weekly morbidity assessment for incidence of diarrhoea and malaria; growth and plasma zinc concentration | Weekley, total 16, 32 or 48 weeks |
| 24 | 10% milk RUTF | Clinics in southern Malawi | Rural | A-S: Children with SAM: WHZ <-3 and or having bipedal pitting oedema. | *Nutritional:*<br>(1) RUTF (25% milk); (2) RUTF (10% milk) | Nutritional recovery | Bi-weekly follow-up visits for up to 4 visits. |
| 25 | Abx for SAM | Clinics in southern Malawi | Rural | A-S: Children with SAM: WHZ <-3 and or having bipedal pitting oedema. | *Nutritional and medical:*<br>(1) 80-90mg/kg/day amoxicillin in 2 daily doses; (2) 14mg/kg/day cefdinir in 2 daily doses; (3) placebo in 2 daily doses.<br><br>All: Counselling + 175kcal/kg/day RUTF | Nutritional recovery | Bi-weekly follow-up visits for up to 6 visits |

| | | | | | | | |
|---|---|---|---|---|---|---|---|
| 26 | 3 foods for MAM | Clinics in southern Malawi | Rural | A-S: Presenting at clinic. Children with MAM (WHZ <-2 and ≥-3 without bipedal oedema) | *Nutritional treatment:* ~75kcal/kg/day<br>(1) CSB++; (2) soy RUSF<br>(3) soy/whey RUSF | Nutritional recovery | Bi-weekly follow-up visits for up to 6 visits |
| 27 | COMPAS | Health facilities and clinics in Kenya and South Sudan | | A-S: Children admitted to an outpatient nutrition program with MUAC <12.5cm and/or oedema. Additional active case finding in the community. | *Nutritional:*<br>SAM: RUTF (200kcal/kg/day).<br>MAM: RUSF (500 kcal/day).<br><br>All SAM: Medical and nutritional evaluation weekly + medication national protocol<br>All MAM: nutritional counselling | Recovery: MUAC ≥ 12.5 cm and no oedema for 2 consecutive visits. | SAM weekly, MAM bi-weekly |
| 28 | DIVIDS | Hospital in study area, India | Urban | A-S: Singleton birth (> 37 weeks gestation), birthweight between 1.8 and 2.5kg, age < 48 hours. | *Nutritional:*<br>(1) Weekly vitamin D supplement for 6 months (35µg/week), (2) placebo | Admission to hospital or death during first 6 months. | Monthly visit, 6 months total. |
| 29 | HI MAM | Clinics, Sierra Leone | | A-S: Children presenting to clinic with uncomplicated MAM (MUAC 11.5-12.4 cm). | *Nutritional and medical:*<br>(1) High risk MAM children (MUAC<11.9 or WAZ<-3.5): 1 packet of RUTF daily and course of amoxicillin until MUAC > 12.4cm or 12 weeks follow-up; low-risk: counselling only<br><br>All: nutritional counselling | Nutritional status 12wk after enrolment. | 6, 12, 24 weeks total. |
| 30 | CHAIN | Hospitals in Kenya, Uganda, Malawi, Burkina Faso, Bangladesh, Pakistan | Rural | I-S: Children admitted with an acute illness, stratified by anthropometric measurements | N/A | 30-day mortality from hospital admission and post-discharge mortality (180 days) | 45 and 90 days post discharge. Up to 180 days plus the duration of admission in total |

| | | | | | | | |
|---|---|---|---|---|---|---|---|
| 31 | SMART 0-6 Phase 1 | Hospitals in Mbarara, Uganda | Rural | I-S: Children admitted to hospital with a proven or suspected infection. | N/A | Post-discharge mortality at any time during 6 months post-discharge period | Every 2 months, 6 months total |
| 32 | SMART 6-60 Phase 0 | Hospitals in Mbarara, Uganda | Rural | I-S: Children admitted to hospital with a proven or suspected infection. | N/A | Post-discharge mortality at any time during 6 months post-discharge period | Every 2 months, 6 months total |
| 33 | SMART6-60 Phase 1 | Hospitals in Mbarara, Uganda | Rural | I-S: Children admitted to hospital with a proven or suspected infection. | N/A | Post-discharge mortality at any time during 6 months post-discharge period | Every 2 months, 6 months total |

* Categorized based on inclusion and exclusion criteria of individual studies. Anthropometry selected (A-S): enrolment based on an anthropometric deficit; Illness selected (I-S): enrolment based on the presence of an illness; General Population (GP): study population enrolled from a general population without consideration of an anthropometric deficit or the presence of illness; other inclusion criteria were however, implemented.

**Table 2:** List of available variables in the WHO Child Mortality Risk Stratification Multi-Country Pooled Cohort (WHO-CMRS)

| Variable/ explanation | Format | Nr of studies |
|---|---|---|
| **Required variables** | | |
| Unique child identifier | Any type/ text | 33 |
| Child’s age at contact (in months) | Continuous (1 decimal) | 33 |
| Child’s sex | Categorical (male/ female) | 33 |
| Did the child die? | Categorical (y/n) | 33 |
| Child’s age at death (in months) | Continuous (1 decimal) | 33 |
| Child's weight (in grams) | Continuous | 33 |
| **Desired / optional variables** | | |
| ***Anthropometry*** | | |
| Child's body length/height (in cm) | Continuous | 33 |
| Mid-upper arm circumference (in mm) | Continuous | 27 |
| Head circumference (in cm) | Continuous | 16 |
| ***Morbidity (definitions as used in respective studies)*** | | |
| Acute diarrhoea (within the last 1, 3, 7, or 14 days) | Categorical (y/n) | 29 |
| Persistent diarrhoea: Any diarrhoea lasting at least 14 days | Categorical (y/n) | 9 |
| Dysentery / Blood in stool in the last 14 days? | Categorical (y/n) | 11 |
| Pneumonia in the last 14 days (diagnosed by physician) | Categorical (y/n) | 5 |
| Pneumonia or acute lower respiratory tract infection (ALRTI) according to WHO definition based on symptoms (fast breathing and cough) in the last 14 days | Categorical (y/n) | 10 |
| Malaria (positive diagnostic test in the last 14 days) | Categorical (y/n) | 11 |
| Positive HIV test for the child (can be in combination with mother's test results in children <18 months) | Categorical (y/n) | 9 |
| Bilateral pitting oedema of feet | Categorical (y/n) | 18 |
| Cough reported by parents in the last 14 days | Categorical (y/n) | 25 |
| Fever reported by parents in the last 3-14 days | Categorical (y/n) | 25 |
| Fever measured by health personnel in the last 14 days | Categorical (y/n) | 8 |
| Acute upper respiratory infection in the last 14 days | Categorical (y/n) | 7 |

| | | |
|---|---|---|
| Grade of oedema according to WHO classification | Categorical (no, mild, moderate, severe) | 3 |
| Haemoglobin concentration in g/dL | Continuous | 13 |
| Currently any anaemia (haemoglobin <11g/dl) | Categorical (y/n) | 13 |
| Severe anaemia (haemoglobin <7 g/dl) | Categorical (y/n) | 13 |
| Does the child have palmar pallor (unusual paleness of the skin)? | Categorical (y/n) | 3 |
| Any hospital admission (overnight) within the last 14 days | Categorical (y/n) | 6 |
| Does the child have a low heart rate? (<100 up to age 1 month; <90 older than 1 month) | Categorical (y/n) | 5 |
| Does the child have leukopenia? (Leukocytes $<6\times10^3$/mm) | Categorical (y/n) | 2 |
| What is the peripheral oxygen saturation? | Continuous (as percent) | 4 |
| Does the child have severe respiratory distress (fast breathing, nasal flaring, wheezing) | Categorical (y/n) | 10 |
| Is the child conscious? | Categorical (no, lethargic, unconscious) | 4 |
| Does the child have jaundice? | Categorical (y/n) | 1 |
| Did the child have seizures or convulsions? | Categorical (y/n) | 9 |
| Is the child able to feed (drink or breastfeed?) | Categorical (y/n) | 12 |
| Does the child show signs of hypotonia? | Categorical (y/n) | 2 |
| Was the child diagnosed with encephalopathy? | Categorical (y/n) | 2 |
| Does the child have sepsis? | Categorical (y/n) | 4 |
| Blood glucose level (in mmol/L) | Continuous | 3 |
| Has the child been vaccinated according to recommended schedule? | Categorical (y/n) | 6 |
| Did the child have any severe underlying morbidity? | Categorical (y/n) | 2 |
| Treatment allocation | Categorical (study arm) | 21 |
| ***Breastfeeding practices (definitions as used in respective studies)*** | | |
| Has the child ever been breastfed? | Categorical (y/n) | 13 |
| Any breastfeeding in the last 14 days **OR**<br>Is the child currently breastfed? | Categorical (y/n) | 26 |
| Is the child exclusively breastfed? | Categorical (no BF, EBF, predominant BF) | 16 |
| Are any fluids or solids (except breastmilk) given (if child <6 months)? | Categorical (y/n) | 11 |

| | | |
|---|---|---|
| ***Birth outcomes*** | | |
| Birthweight (in grams) | Continuous | 17 |
| Gestational age at birth (in completed weeks) * | Continuous | 13 |
| ***Socio-demographic and parental characteristics*** | | |
| Child’s date of birth | Date: MM/YYYY | 29 |
| Maternal age at enrolment (in years) | Continuous | 25 |
| Is the mother alive? | Categorical (y/n) | 21 |
| Is the father alive? | Categorical (y/n) | 11 |
| Is the biological mother the primary caregiver? | Categorical (y/n) | 14 |
| Years of formal education of the mother | Preferably continuous | 20 |
| Is the mother literate? | Categorical (y/n) | 16 |
| Years of formal education of the father | Preferably continuous | 12 |
| Is the father literate? | Categorical (y/n) | 10 |
| Mother's body mass index (BMI in $kg/m^2$) | Continuous (1 decimal) | 19 |
| Mother diagnosed with tuberculosis? | Categorical (y/n) | 3 |
| Maternal HIV status | Categorical (negative/ positive) | 10 |
| ***Study and population level characteristics*** | | |
| Improved drinking water sources available in the household? (WHO definition) | Categorical (y/n) | 15 |
| Improved sanitation facilities available in the household? (WHO definition) | Categorical (y/n) | 13 |
| Did the child die in a health facility or at home? | Categorical (facility, home) | 6 |

**Ascertained by date of last menstrual period, early ultrasound or study specific algorithm*

### Table S4: Availability of variables in individual studies included in the WHO Child Mortality Risk Stratification Multi-Country Pooled Cohort (WHO CMRS)

| | 1 AMANHI Bangladesh | 2 AMANHI Pakistan | 3 CARING | 4 CHILD2 | 5 CRI study | 6 IECDZ study | 7 iLiNS ZINC | 8 Lao Zinc | 9 Low BW | 10 MAL-ED Bangladesh | 11 MAL-ED India | 12 MISAME I | 13 MISAME II | 14 MISAME III | 15 MRCG Keneba | 16 Pemba | 17 PM2A Guatemala | 18 PNS | 19 PROMIS Burkina | 20 PROMIS Mali | 21 ROSE | 22 South Africa study | 23 ZINC720 | 24 3 foods for MAM | 25 10% milk RUTF | 26 Abx for SAM | 27 ComPAS | 28 DIVIDS | 29 HI MAM | 30 CHAIN | 31 SMART 0-6 Phase 1 | 32 SMART 6-60 Phase 0 | 33 SMART 6-60 Phase 1 | **Total** |
|---|---|---|---|---|---|---|---|---|---|---|---|---|---|---|---|---|---|---|---|---|---|---|---|---|---|---|---|---|---|---|---|---|---|---|
| **Required variables** | | | | | | | | | | | | | | | | | | | | | | | | | | | | | | | | | | |
| Unique child identifier | X | X | X | X | X | X | X | X | X | X | X | X | X | X | X | X | X | X | X | X | X | X | X | X | X | X | X | X | X | X | X | X | X | 33 |
| Child's age at contact (in months) | X | X | X | X | X | X | X | X | X | X | X | X | X | X | X | X | X | X | X | X | X | X | X | X | X | X | X | X | X | X | X | X | X | 33 |
| Child's sex | X | X | X | X | X | X | X | X | X | X | X | X | X | X | X | X | X | X | X | X | X | X | X | X | X | X | X | X | X | X | X | X | X | 33 |
| Did the child die? | X | X | X | X | X | X | X | X | X | X | X | X | X | X | X | X | X | X | X | X | X | X | X | X | X | X | X | X | X | X | X | X | X | 33 |
| Child's age at death (in months) | X | X | X | X | X | X | X | X | X | X | X | X | X | X | X | X | X | X | X | X | X | X | X | X | X | X | X | X | X | X | X | X | X | 33 |
| Child's weight (in grams) | X | X | X | X | X | X | X | X | X | X | X | X | X | X | X | X | X | X | X | X | X | X | X | X | X | X | X | X | X | X | X | X | X | 33 |
| **Desired / optional variables** | | | | | | | | | | | | | | | | | | | | | | | | | | | | | | | | | | |
| ***Anthropometry*** | | | | | | | | | | | | | | | | | | | | | | | | | | | | | | | | | | |
| Child's body length/height (in cm) | X | X | X | X | X | X | X | X | X | X | X | X | X | X | X | X | X | X | X | X | X | X | X | X | X | X | X | X | X | X | X | X | X | 33 |
| Mid-upper arm circumference (in mm) | X | X | X | X | | | X | X | X | | | X | X | X | X | X | | X | X | X | X | | X | X | X | X | X | X | X | X | X | X | X | 27 |
| Head circumference (in cm) | X | X | | X | | | X | | X | X | X | X | X | X | X | X | | X | | | X | | | | | | | X | | X | | | | 16 |
| ***Morbidity (definitions as used in respective studies)*** | | | | | | | | | | | | | | | | | | | | | | | | | | | | | | | | | | |
| Acute diarrhoea (within the last 1, 3, 7, or 14 days) | | | X | X | X | X | X | X | X | X | X | X | X | X | | | X | X | X | X | X | X | X | X | X | X | X | X | X | X | X | X | X | 29 |
| Persistent diarrhoea: Any diarrhoea lasting at least 14 days | | | | X | | | X | X | | | | | | | | | | X | | | X | | X | | | | | | | X | X | | X | 9 |
| Dysentery / Blood in stool in the last 14 days? | | | X | X | | | X | X | | X | | | | | | | X | | X | X | X | | X | | | | | | | X | | | | 11 |
| Pneumonia in the last 14 days (diagnosed by physician) | | | | X | | | X | X | | X | X | | | | | | | X | X | X | | | X | | | | | | | X | X | X | X | 13 |
| Malaria (positive diagnostic test in the last 14 days) | | | | X | | | X | | | | | | | | | | | X | X | X | X | | X | | | | | | | X | X | X | X | 11 |
| Positive HIV test for the child | | | | X | | | | | | | | | | | | | | | | | | | | X | | X | X | | X | X | X | X | X | 9 |

| | | | | | | | | | | | | | | | | | | | | | | | | | | | | | | | | | | |
|---|---|---|---|---|---|---|---|---|---|---|---|---|---|---|---|---|---|---|---|---|---|---|---|---|---|---|---|---|---|---|---|---|---|---|
| Bilateral pitting oedema of feet | | | X | X | | | X | | X | X | | | | X | | | | | X | X | X | | X | X | X | X | X | X | X | X | X | | | 18 |
| Cough reported by parents in the last 14 days | | | X | | | X | X | X | X | X | | X | X | X | | | X | X | X | X | X | X | X | X | X | X | X | X | X | X | X | | X | 25 |
| Fever reported by parents in the last 3-14 days | | | X | | | X | X | X | X | X | | X | X | X | | | X | X | X | X | X | X | X | X | X | X | X | X | X | X | X | | X | 25 |
| Fever measured by health personnel in the last 14 days | | | | X | | | X | X | | | | | | X | | | | | X | X | | | X | | | | | | | X | | | | 8 |
| Acute upper respiratory infection in the last 14 days | | | | X | | | X | X | | | | | | | | | | | | | | | X | | | | | | | X | X | | X | 7 |
| Grade of oedema according to WHO classification | | | | | | | | | | | | | | | | | | | | | | | | | | | X | | X | X | | | | 3 |
| Haemoglobin concentration in g/dL | | | | X | | | X | X | | X | | | | X | | | X | X | | | X | | X | | | | | | | X | X | X | X | 13 |
| Currently any anaemia (haemoglobin <11g/dl) | | | | X | | | X | X | | X | | | | X | | | X | X | | | X | | X | | | | | | | X | X | X | X | 13 |
| Severe anaemia (haemoglobin <7 g/dl) | | | | X | | | X | X | | X | | | | X | | | X | X | | | X | | X | | | | | | | X | X | X | X | 13 |
| Does the child have palmar pallor (unusual paleness of the skin)? | | | | X | | | | | | | | | | | | | | X | | | | | | | | | | | | | X | | | 3 |
| Any hospital admission (overnight) within the last 14 days | | | | | | | | | | | | | | | | | | | | | X | | X | | | | X | | | X | X | X | | 6 |
| Does the child have a low heart rate? (<100 up to age 1 month; <90 older than 1 month) | | | | | | | | | | | | | | | | | | | | | X | | | | | | | | | X | X | X | X | 5 |
| Does the child have leukopenia? (Leukocytes <6×10³/mm) | | | | | | | | | | | | | | | | | | | | | | | | | | | | | | X | | X | | 2 |
| What is the peripheral oxygen saturation? | | | | | | | | | | | | | | | | | | | | | | | | | | | | | | X | X | X | X | 4 |
| Does the child have severe respiratory distress (fast breathing, nasal flaring, wheezing) | | | | X | | | X | X | | | | | | | | | | | X | X | X | | X | | | | | | | X | X | | X | 10 |
| Is the child conscious? | | | | X | X | | | | | | | | | | | | | | | | | | X | | | | | | | X | | | | 4 |
| Does the child have jaundice? | | | | | | | | | | | | | | | | | | | | | | | | | | | | | | X | | | | 1 |
| Did the child have seizures or convulsions? | | | | X | | | X | X | | | | | | | | | | | X | X | | | X | | | | | | | X | X | | X | 9 |

| | | | | | | | | | | | | | | | | | | | | | | | | | | | | | | | | | | |
|---|---|---|---|---|---|---|---|---|---|---|---|---|---|---|---|---|---|---|---|---|---|---|---|---|---|---|---|---|---|---|---|---|---|---|
| Is the child able to feed (drink or breastfeed?) | | | | | | | X | X | | | | | | | | | | X | X | X | | | X | X | X | X | | | | X | X | | X | 12 |
| Does the child show signs of hypotonia? | | | | | | | | | | | | | | | | | | | | | | | | | | | | | | X | X | | | 2 |
| Was the child diagnosed with encephalopathy? | | | | X | | | | | | | | | | | | | | | | | | | | | | | | | | X | | | | 2 |
| Does the child have sepsis? | | | | | | | | | | | | | | | | | | | | | | | | | | | | | | X | X | X | X | 4 |
| Blood glucose level (in mmol/L) | | | | | | | | | | | | | | | | | | | | | | | | | | | | | | X | X | | X | 3 |
| Has the child been vaccinated according to recommended schedule? | | X | X | | | | X | | | | | | | | | | | | X | | X | | X | | | | | | | | | | | 6 |
| Did the child have any severe underlying morbidity? | | | | | | | | | | | | | | | | | | | | | | | | | | | X | | | X | | | | 2 |
| Treatment allocation | | | X | X | | X | X | X | | | | X | X | X | | | X | X | X | X | X | X | X | X | X | X | X | X | X | | | | | 21 |
| ***Breastfeeding practices (definitions as used in respective studies)*** | | | | | | | | | | | | | | | | | | | | | | | | | | | | | | | | | | |
| Has the child ever been breastfed? | | X | X | X | X | | | | X | | X | | | X | | | X | | | | X | | | | | | X | | | | X | X | X | 13 |
| Any breastfeeding in the last 14 days | | | X | X | X | X | X | X | X | X | X | | | | | | X | X | X | X | X | X | X | X | X | X | X | X | X | X | X | X | X | 26 |
| Is the child exclusively breastfed? | | X | X | X | X | | | | X | X | X | | | X | | | X | X | X | | X | | | | | | | X | | X | X | | X | 16 |
| Are any fluids or solids (except breastmilk) given (if child <6 months)? | | X | X | X | X | | | | | X | X | | | | | | X | | X | | X | | | | | | | | | X | X | | | 11 |
| ***Birth outcomes*** | | | | | | | | | | | | | | | | | | | | | | | | | | | | | | | | | | |
| Birthweight (in grams) | X | X | X | X | X | | | | X | X | X | X | X | X | | X | | X | | | X | X | | | | | | | X | X | | | | 17 |
| Gestational age at birth (in completed weeks) * | X | X | | X | | | | | X | | | X | X | X | | X | X | X | | | X | | | | | | | | | | X | | X | 13 |
| ***Socio-demographic and parental characteristics*** | | | | | | | | | | | | | | | | | | | | | | | | | | | | | | | | | | |
| Child's date of birth | X | X | X | X | X | X | X | X | X | X | X | X | X | | X | X | X | X | X | X | X | X | X | X | X | X | X | X | X | X | | | | 29 |
| Maternal age at enrolment (in years) | X | X | X | X | X | X | X | X | X | X | X | X | X | X | | X | X | X | X | X | X | | | | | | | X | | X | X | X | X | 25 |
| Is the mother alive? | | X | X | X | | X | X | X | X | X | X | | | | | | X | | | | X | X | | X | X | X | X | | X | X | X | X | X | 21 |
| Is the father alive? | | | | X | | X | X | | | X | | | | | | | | | | | | X | | X | X | X | X | | X | X | | | | 11 |
| Is the biological mother the primary caregiver? | | | | | | | X | X | X | | | | | | | | X | | X | X | | | | X | | X | X | X | X | X | X | | X | 14 |
| Years of formal education of the mother | X | X | X | X | | | X | X | X | | | | | X | | X | X | X | X | X | X | | | | | | X | X | | X | X | X | X | 20 |
| Is the mother literate? | | X | X | X | | | X | X | X | X | | X | X | | | | X | | X | X | X | | X | | | | | X | | X | | | | 16 |

| | | | | | | | | | | | | | | | | | | | | | | | | | | | | | | | | | | |
|---|---|---|---|---|---|---|---|---|---|---|---|---|---|---|---|---|---|---|---|---|---|---|---|---|---|---|---|---|---|---|---|---|---|---|
| Years of formal education of the father | X | X | | X | | | X | | X | | | | | | | | X | X | X | X | X | | | | | | X | X | | | | | | 12 |
| Is the father literate? | | X | | X | | | X | | X | | | X | X | | | | | | X | X | X | | | | | | | X | | | | | | 10 |
| Mother's body mass index (BMI in kg/m²) | X | X | X | | | | X | X | X | X | | X | X | X | | X | X | X | X | X | X | | X | | | | | X | | X | | | | 19 |
| Mother diagnosed with tuberculosis? | | | | | | | | | | | | | | | | | | X | | | | | | | X | X | | | | | | | | 3 |
| Maternal HIV status | | | | X | | X | | | | | | | | | | | | X | | | | | | X | | X | | | X | X | X | X | X | 10 |
| ***Study and population level characteristics*** | | | | | | | | | | | | | | | | | | | | | | | | | | | | | | | | | | |
| Improved drinking water sources available in the household? (WHO definition) | | X | X | | | | X | X | | | | | | X | | | X | | X | X | X | | X | | | | X | | | X | X | X | X | 15 |
| Improved sanitation facilities available in the household? (WHO definition) | | X | X | | | | X | X | | | | | | X | | | X | | X | X | X | | X | | | | X | | | X | | | | 12 |
| Did the child die in a health facility or at home? | | | | | | | | | | | | | | | | | | | | | X | | X | | | | | | | X | X | X | X | 6 |

**Table S5: Selected maternal and child characteristics of individual studies included in the WHO Child Mortality Risk Stratification Multi-Country Pooled Cohort (WHO CMRS)**

| Study Nr | Short name | Category* (GP; AS; IS) | Nr of children | Nr of deaths | Median (IQR) total FU time (mo) | Median FU frequency (mo) | Sex (male) | Age (months) at enrolment, median (IQR) | Prevalence wasting at enrolment (WHZ <-2) | Prevalence underweight at enrolment (WAZ <-2) | Prevalence stunting at enrolment (HAZ <-2) | Proportion mothers without education |
|---|---|---|---|---|---|---|---|---|---|---|---|---|
| 1 | AMANHI Bangladesh | GP | 938 | 44 | 23.9 (23.8, 24) | 5.6 (3.2, 6.0) | 49% | 0.1 (0.1, 0.2) | 12.8 | 28.4 | 32.0 | 3.9 |
| 2 | AMANHI Pakistan | GP | 1,138 | 39 | 23.7 (23.6, 23.8) | 5.7 (2.9, 6.1) | 50% | 0.1 (0, 0.1) | 12.2 | 21.1 | 21.6 | 48.1 |
| 3 | CARING | GP | 2,993 | 63 | 17.7 (17.5, 17.7) | 3 (2.9, 3.2) | 50% | 0.1 (0.1, 0.2) | 16.8 | 36.2 | 43.4 | 54.7 |
| 4 | CHILD 2 | GP | 2,398 | 45 | 17.5 (14.3, 17.7) | 0.9 (0.7, 1.0) | 51% | 1.3 (1.3, 1.4) | 5.9 | 3.9 | 7.4 | 1.6 |
| 5 | CRI | GP | 373 | 0 | 43.9 (40.4, 47.7) | 1 (0.8, 1.1) | 50% | 0 (0, 0) | 12.9 | 11.5 | 17.8 | N/A |
| 6 | IECDZ Study | GP | 526 | 8 | 27.2 (26.7, 27.6) | 13.4 (12.7, 14.6) | 50% | 8.85 (7, 10.9) | 6.8 | 13.9 | 43.7 | N/A |
| 7 | iLiNS-Zinc | GP | 3,220 | 58 | 8.9 (8.7, 9.0) | 3 (2.8, 3.2) | 51% | 9.4 (9.1, 9.7) | 16.4 | 29.5 | 22.5 | 85.8 |
| 8 | Lao zinc | GP | 3,406 | 4 | 7.8 (7.7, 7.9) | 0.9 (0.7, 1.0) | 51% | 13.7 (9.9, 18.6) | 8.0 | 26.7 | 39.1 | 20.1 |
| 9 | Low BW | GP | 1,103 | 86 | 12 (12, 12) | 1 (1, 1) | 52% | 0 (0,0) | 31.3 | 16.8 | 10.3 | 76.8 |
| 10 | MAL-ED Bangladesh | GP | 265 | 3 | 35 (35, 35) | 1 (1, 1.1) | 50% | 0.99 (0.95, 1.02) | 3.6 | 12.7 | 19.0 | N/A |
| 11 | MAL-ED India | GP | 247 | 1 | 35.1 (28.1, 35.1) | 1 (1, 1) | 46% | 1.1 (1.1, 1.1) | 9.3 | 29.2 | 23.9 | N/A |
| 12 | MISAME I | GP | 1,221 | 104 | 12 (11.2, 12.4) | 1.2 (1.0, 1.8) | 51% | 0 (0,0) | 12.1 | 13.6 | 14.4 | N/A |
| 13 | MISAME II | GP | 1,084 | 55 | 12.1 (11.5, 12.5) | 1.1 (1.0, 1.6) | 51% | 0 (0,0) | 7.5 | 12.7 | 18.1 | N/A |
| 14 | MISAME III | GP | 1,608 | 72 | 10.8 (8.0, 11.1) | 1.1 (0.9, 2.1) | 49% | 1.05 (0.95, 1.14) | 3.0 | 7.1 | 11.7 | 58.4 |
| 15 | MRCG Keneba | GP | 560 | 9 | 17.7 (11.7, 21.9) | 1.2 (0.7, 2.8) | 53% | 1.8 (0.6, 7.3) | 4.1 | 10.7 | 16.1 | N/A |
| 16 | Pemba | GP | 4,109 | 99 | 36.1 (35.4, 37.2) | 5.9 (4.6, 7) | 51% | 0 (0,0) | 3.2 | 5.4 | 10.7 | 13.9 |
| 17 | PM2A | GP | 4,212 | 67 | 22.9 (22.7, 23.0) | 3 (2.8, 5.9) | 49% | 1.05 (0.5, 1.24) | 2.3 | 9.1 | 17.5 | 33.2 |
| 18 | PNS | GP | 7,956 | 297 | 15.4 (11.4, 16.9) | 0.9 (0.9, 1.0) | 51% | 0 (0, 0.1) | 5.0 | 9.9 | 16.2 | 7.1 |
| 19 | PROMIS Burkina Faso | GP | 2,113 | 50 | 17.7 (17.3, 18.1) | 1 (0.9, 1.1) | 51% | 0.63 (0.40, 0.90) | 7.6 | 6.4 | 5.9 | 83.8 |

| | | | | | | | | | | | | |
|---|---|---|---|---|---|---|---|---|---|---|---|---|
| 20 | PROMIS Mali | GP | 1,132 | 36 | 18 (18, 18.1) | 1 (1, 1) | 52% | 6.51 (6.26, 6.76) | 0 | 10.3 | 11.5 | 79.5 |
| 21 | ROSE | GP | 2,507 | 36 | 23.8 (23.3, 24.3) | 0.9 (0.7, 1.2) | 51% | 0.47 (0, 0.7) | 7.1 | 24.7 | 36.4 | 28.8 |
| 22 | South Africa Study | GP | 386 | 0 | 27.7 (22.5, 30.6) | 10.3 (8.9, 21.1) | 47% | 7.69 (7.03, 9.04) | 1.0 | 2.1 | 8.8 | N/A |
| 23 | ZINC 720 | GP | 6,245 | 62 | 3.8 (3.4, 7.3) | 3.7 (3.6, 3.8) | 51% | 13.8 (9, 20) | 14.3 | 28.7 | 29.5 | N/A |
| 24 | 3 foods for MAM | A-S | 2,707 | 25 | 1.4 (0.5, 3) | 0.5 (0.5, 0.5) | 36% | 16.4 (11.4, 24.3) | 76.7 | 93.2 | 72.3 | N/A |
| 25 | 10% milk RUTF | A-S | 1,873 | 64 | 1.0 (0.5, 1.8) | 0.5 (0.4, 0.5 | 43% | 17.1 (12, 24.2) | 51.7 | 80.3 | 75.9 | N/A |
| 26 | Abx for SAM | A-S | 2,742 | 147 | 1.0 (0.5, 2) | 0.5 (0.5, 0.5) | 46% | 19.4 (13.4, 26.8) | 48.4 | 78.4 | 80.3 | N/A |
| 27 | COMPAS | A-S | 4,072 | 35 | 3.0 (1.8, 3.9) | 0.5 (0.2, 0.5) | 41% | 13 (9, 24) | 66.6 | 80.3 | 52.1 | 53.2 |
| 28 | DIVIDS | A-S | 2,079 | 39 | 6.0 (5.9, 6.0) | 0.9 (0.9, 1.1) | 47% | 0 (0, 0) | 44.0 | 89.2 | 51.2 | 19.5 |
| 29 | HI MAM | A-S | 1,286 | 56 | 5.5 (5.5, 5.5) | 0.5 (0.5, 1.8) | 41% | 11.5 (8.3, 16.6) | 36.7 | 87.6 | 74.5 | N/A |
| 30 | CHAIN | I-S | 3,101 | 350 | 6.1 (5.9, 6.4) | 1.7 (1.5, 2.8) | 57% | 10.7 (6.8, 15.6) | 49.9 | 62.4 | 49.8 | 26.3 |
| 31 | SMART 0-6 Phase 1 | I-S | 2,647 | 384 | 6 (6, 6) | 6 (6, 6) | 56% | 1.5 (0.4, 3.6) | 28.4 | 25.9 | 23.8 | 5.9 |
| 32 | SMART 6-60 Phase 0 | I-S | 1,273 | 130 | 6 (6, 6) | 6 (6, 6) | 55% | 18.1 (10.9, 34.4) | 34.7 | 28.4 | 25.6 | 28.0 |
| 33 | SMART6-60 Phase 1 | I-S | 3,767 | 337 | 6 (6, 6) | 6 (6, 6) | 55% | 16.8 (10.8, 28.1) | 29.2 | 30.2 | 27.2 | 7.7 |
| **ALL** | | | **75,287** | **2,805** | **17 (8, 23)** | **1 (0.9, 1.5)** | **49.8%** | **2.9 (0.1, 12.3)** | **24.1** | **35.5** | **33.3** | **33.4** |

* Categorized based on inclusion and exclusion criteria of individual studies. Anthropometry selected (A-S): enrolment based on an anthropometric deficit; Illness selected (I-S): enrolment based on the presence of an illness; General Population (GP): study population enrolled from a general population without consideration of an anthropometric deficit or the presence of illness; other inclusion criteria were however, implemented

**Table S6a** – Maternal and child characteristics in the WHO Child Mortality Risk Stratification Multi-Country Pooled Cohort (WHO CMRS) according to study population category

| Characteristic | GP[1] | A-S[1] | I-S[1] |
|---|---|---|---|
| ***Maternal characteristics at enrolment*** | *% or median (n[3])* | *% or median (n[3])* | *% or median (n[3])* |
| Mother alive | 99.8% (19,661) | 98 (12,247) | 99 (10,611) |
| Mother primary caretaker | 98% (14,764) | 95 (12,176) | 94 (8,869) |
| Median maternal age, years | 25 years | 23 years | 26 years |
| Maternal education | | | |
| no education | 34% (12,284) | 37 (2,207) | 17 (1,243) |
| primary education | 25% (8,880) | 10 (578) | 25 (1,848) |
| secondary or higher education | 41% (14,475) | 53 (3,156) | 58 (4,381) |
| Mother HIV positive | 0.2 (17) | 15 (644) | 18 (280) |
| ***Child's characteristics at enrolment*** | *% or median (n[2])* | *% or median (n[2])* | *% or median (n[2])* |
| Child's sex, male | 51 (25,133) | 43 (6,313) | 56 (6,001) |
| Child's age, months<br>0 | 30 (15,212) | 14 (2,079) | 1 (92) |
| 1-5 | 39 (19,211) | 0.04 (6) | 30 (3,192) |
| 6-11 | 19 (9,200) | 28 (4,193) | 25 (2,663) |
| 12-23 | 11 (5,539) | 35 (5,194) | 29 (3,146) |
| 24-59 | 1 (577) | 22 (3,287) | 16 (1,695) |
| Low birthweight, <2500g | 13 (3,536) | 5 (55) | 21 (452) |
| Median birthweight, kg | 3 (2.7, 3.4) | 3 (2.9, 3,4) | 3 (2.5, 3.5) |
| Preterm birth, <37 weeks of gestation | 16 (4,353) | No data | 4 (274) |
| Median (IQR) WAZ | -0.9 (-1.7, -0.1) | -2.9 (-3.6, -2.3) | -1.4 (-2.8, -0.4) |
| Median (IQR) HAZ | -1.1 (-1.9, -0.3) | -2.5 (-3.5, -1.7) | -1.2 (-2.4, 0.1) |
| Median (IQR) WHZ | -0.3 (-1.2, 0.6) | -2.2 (-2.8, -1.6) | -1.3 (-2.6, 0.1) |
| Median (IQR) MUAC | 120 (104, 135) | 118 (110, 123) | 128 (114, 141) |
| ***Child's characteristics at any visit*** | *% or median (n[3])* | *% or median (n[3])* | *% or median (n[3])* |
| Child's age, months | | | |
| 0-1 | 10 (47,493) | 6 (3,896) | 8 (1,483) |
| 2-5 | 20 (94,566) | 11 (6,872) | 11 (2,142) |
| 6-11 | 29 (135,464) | 26 (16,734) | 27 (4,999) |
| 12-17 | 21 (97,325) | 22 (14,569) | 25 (4,645) |
| 18-23 | 12 (52,907) | 13 (8,521( | 16 (3,031) |
| 24-59 | 8 (35,450) | 22 (13,929) | 13 (2,434) |
| Acute diarrhea[4] | 8 (23,730) | 30 (16,108) | 44 (8,160) |
| Acute lower respiratory tract infection[4] | 3 (4,844) | No data | 40 (7,484) |
| Severe illness[4] | 1.2 (2,917) | 10 (3,483) | 28 (4,950) |
| Any breastfeeding[4] at any visit | | | |
| at age 0-5 months | 99 (91,031) | 99 (7,854) | 91 (3,303) |
| at age 6-23 months | 87 (176,213) | 82 (20,667) | 71 (8,951) |
| at age 24-59 months | 19 (4,015) | 16 (1,204) | 17 (415) |

[1] Study population type: A-S (anthropometry selected), I-S (illness-selected), and GP (General population: enrolment not based on anthropometry or illness)

[2] Number of children

[3] Number of observations

[4] Study definition used. For details see method-section of the main text.

**Table S6b** – Time varying child characteristics in the WHO Risk Stratification Multi-Country Pooled Cohort (WRS-MCPC) according to age group

| Characteristic | <6 mo | 6-11 mo | 12-23 mo | 24-59 mo |
|---|---|---|---|---|
| ***Child's characteristics at any visit*** | *% (n[1])* | *% (n[1])* | *% (n[1])* | *% (n[1])* |
| Median (IQR) WAZ | -0.7 (-1.8, 0.0) | -1.7 (-2.7, -0.7) | -2.1 (-3.1, -1.2) | -2.6 (-3.6, -1.4) |
| Median (IQR) HAZ | -1.0 (-1.8, -0.1) | -1.4 (-2.3, -0.5) | -2.1 (-3.1, -1.2) | -2.4 (-3.6, -1.4) |
| Median (IQR) WHZ | -0.1 (-1.2, 0.9) | -1.2 (-2.1, -0.3) | -1.6 (-2.4, -0.6) | -1.9 (-2.7, -0.7) |
| Median (IQR) MUAC, mm | 105 (96, 116) | 128 (119, 139) | 129 (120, 140) | 130 (120, 145) |
| Acute diarrhea[2] | 6 (5,946) | 15 (16,301) | 14 (19,721) | 15 (6,029) |
| Acute lower respiratory tract infection[2] | 8 (3,056) | 9 (4,480) | 6 (4,088) | 5 (702) |
| Severe illness[2] | 2 (1,213) | 4 (3,767) | 4 (4,745) | 7 (1,625) |
| Any breastfeeding[2] | 99 (102,188) | 96 (107,520) | 77 (98,311) | 19 (5,934) |

[1] Number of observations

[2]Study definition used. For details see method-section of the main text.

Abbreviations: HAZ = height-for-age Z-score; mo = months; MUAC= mid-upper arm circumference (in mm); WAZ= weight-for-age Z-score; WHZ= weight-for-height Z-score.

**Figure S1**: Mean anthropometric status for children in the in the WHO Risk Stratification Multi-Country Pooled Cohort (WRS-MCPC) by 3-months age group and study population type; GP (not enrolled by anthropometric deficit or illness), A-S (anthropometry-selected), and I-S (illness-selected)

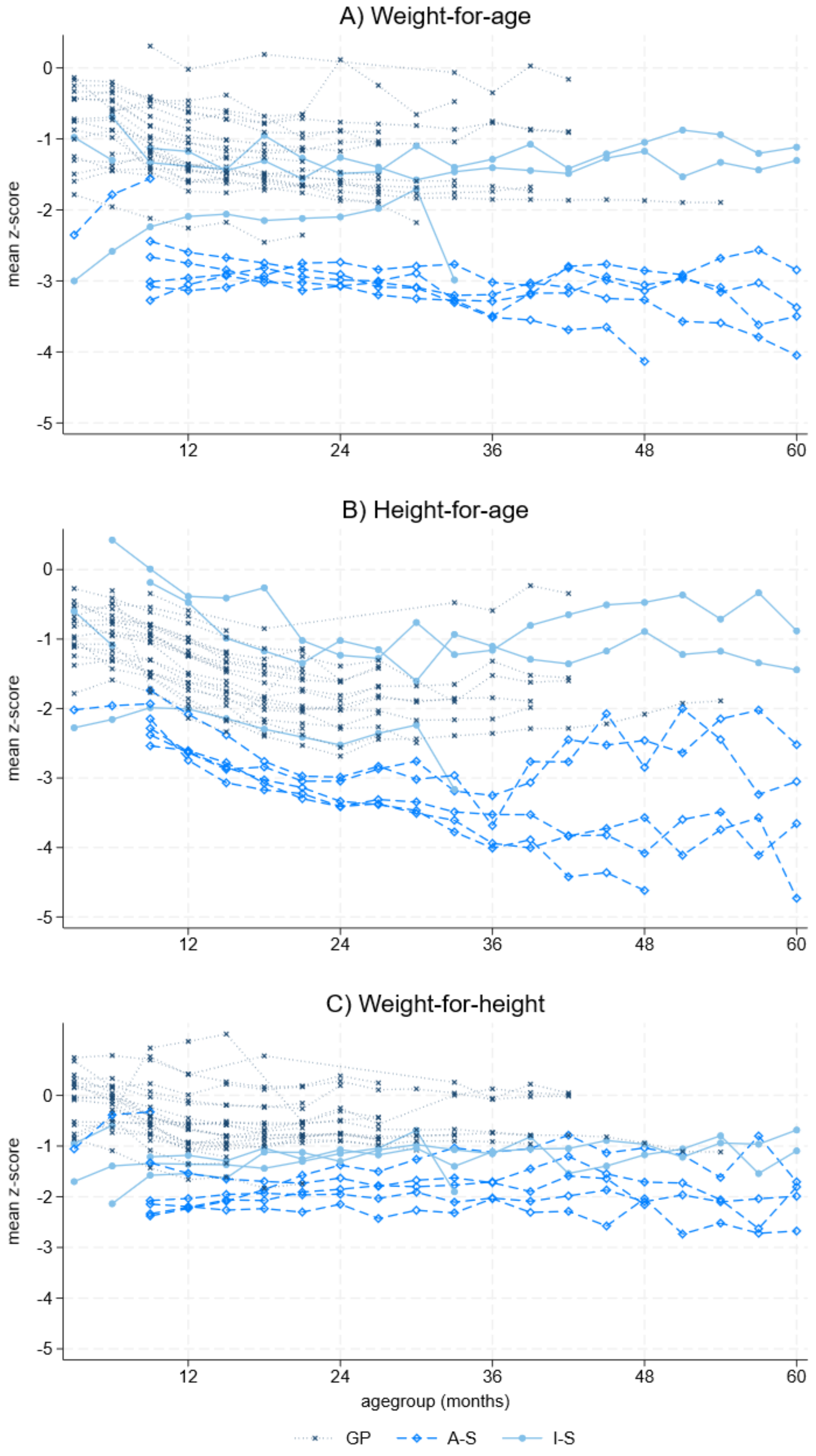
